# Beyond Tumors: Reduced survival linked to pathogenic *PIK3CA* and *TP53* post-zygotic variants in the uninvolved breast tissue of recurrent cancer patients

**DOI:** 10.1101/2024.10.04.24313634

**Authors:** Maria Andreou, Katarzyna Chojnowska, Natalia Filipowicz, Monika Horbacz, Piotr Madanecki, Katarzyna Duzowska, Urszula Ławrynowicz, Hanna Davies, Bożena Bruhn-Olszewska, Mikołaj Koszyński, Kinga Drężek-Chyła, Maciej Jaśkiewicz, Marcin Jąkalski, Anna Kostecka, Marta Drzewiecka-Kłysz, Magdalena Nowikiewicz, Manuela Las-Jankowska, Dariusz Bała, Jacek Hoffman, Ewa Śrutek, Michał Jankowski, Jerzy Jankau, Diana Hodorowicz-Zaniewska, Joanna Szpor, Łukasz Szylberg, Wojciech Zegarski, Tomasz Nowikiewicz, Patrick G. Buckley, Irene Tiemann-Boege, Jakub Mieczkowski, Magdalena Koczkowska, Jan P. Dumanski, Arkadiusz Piotrowski

**Author notes:** A.P., J.P.D, and M. Koczkowska contributed equally. Correspondence: Arkadiusz Piotrowski, 3P-Medicine Laboratory, Medical University of Gdańsk, Gdańsk, Poland.

## Abstract

Histologically normal mammary tissue from breast cancer patients can harbor significant genetic alterations. We analyzed the spectrum of DNA variants in 408 matched histologically normal tissue and tumors from 77 poor-prognosis patients, 49 patients recruited without prognosis bias, and mammary gland samples from 15 non-cancerous individuals. Whole exome sequencing revealed a higher prevalence of pathogenic post-zygotic variants in cancer-associated genes: *AKT1*, *PIK3CA*, *PTEN*, *TBX3*, and *TP53*, affecting poor-prognosis patients (29%), compared to 12.5% in those without prognosis criteria (p=0.0008578). *PIK3CA* variants were recurrent across patients, while *TP53* variants were restricted to those with adverse prognoses. Duplex sequencing detected low-frequency pathogenic *PIK3CA* and *TP53* variants in distant normal tissues of poor-prognosis patients. Disease recurrence significantly reduced survival rates, with poor prognosis patients experiencing higher mortality within 24 months (p=0.0088), further worsened by the presence of pathogenic post-zygotic variants. These findings highlight the importance of genetic monitoring even in microscopically normal mammary tissue.

## INTRODUCTION

Breast cancer accounts for 12.5% of global annual cancer diagnoses, with the incidence rate increasing by 0.5% annually in recent years^1,2^ Despite an overall 42% reduction between 1989 and 2021, primarily attributed to increased awareness and early detection, breast cancer still constitutes one of the leading causes of death among women^3^. Notably, stage I, low-risk breast cancer cases still present a 15-20% chance of recurrence even two decades after the initial diagnosis^4^. While 5-10% of breast cancer cases are hereditary, with 25-30% of heritable breast cancer risk attributed to pathogenic variants in genes of high and moderate penetrance^5^, the majority of cases are considered sporadic^6,7^.

Research has recently focused on the normal mammary gland within the affected breast for early detection of tumor formation at the molecular level, preceding changes on imaging or palpative screens^8^. Concurrently, breast-conserving surgery (BCS), which aims to remove the tumor and preserve the remaining healthy breast tissue, stands as the preferred approach^9,10^. However, mammary gland tissue from breast cancer patients, although appearing normal, has been found to harbor significant genomic and transcriptomic alterations^11–14^. In particular, non-tumorous tissue from patients undergoing BCS shows clearly pathogenic low-level post-zygotic alterations in the *PIK3CA* and *TP53* genes, raising questions regarding its oncogenic potential ^15,16^.

Nevertheless, the association between post-zygotic alterations in ostensibly normal mammary gland tissue of breast cancer patients and their clinical utility remains unclear. Hence, we screened paired uninvolved mammary gland (UM) and primary tumor (PT) samples of reportedly sporadic breast cancer patients with adverse outcomes within 10 years post-original surgery for the presence of post-zygotic alterations. We compared our findings with a second breast cancer cohort of reportedly sporadic patients recruited without specific prognosis criteria. Additionally, normal mammary gland samples from individuals with no personal or family history of cancer were included as controls (study overview in Supplementary Figure 1).

Here, we demonstrate that pathogenic post-zygotic variants in cancer-associated genes are commonly found in histologically normal mammary tissue of breast cancer patients with adverse prognoses. These variants, often also found in corresponding primary tumors, are linked with patient survival, emphasizing the need for molecular screening to improve the clinical management of patients.

## MATERIALS & METHODS

### Patient recruitment, sample collection, and DNA isolation

We carried out Whole Exome Sequencing (WES) on 408 samples from three distinct groups: two cohorts of female breast cancer patients with differing prognostic outcomes and a control group composed of female individuals who underwent mammoplasty surgery for non-cancer-related reasons. The assignment of patients to the cohorts was determined by their clinical prognoses. The first cohort included 77 reportedly sporadic breast cancer patients with adverse outcomes. All individuals in this group experienced either recurrent disease, such as local recurrence/metastasis to the breast or secondary organs (n=40), developed a second independent tumor (n=18), or both (n=8), and/or succumbed to the disease (n=45) within the proceeding 10 years (<u>B</u>reast <u>C</u>ancer <u>A</u>dverse <u>P</u>rognoses cohort, BCAP), (Table 1, Supplementary Table 1). The second cohort included 49 individuals from the same ethnic population, diagnosed with reported sporadic breast cancer but recruited without specific criteria related to prognosis (<u>B</u>reast <u>C</u>ancer <u>U</u>n-<u>S</u>elected cohort, BCUS). Within this group, 5 out of 49 patients experienced recurrence, and 3 of them died within 2 years post-surgery; however, the follow-up period for this cohort was considerably shorter compared to the BCAP cohort (Table 1, Supplementary Table 1). The majority of BCAP and BCUS patients were treated with BCS (n=63 and n=31 respectively) versus mastectomy (n=12 and n=18 respectively) (Supplementary Table 1) (data missing for 2 BCAP patients). All recruited individuals did not receive neoadjuvant therapy. The control group comprised 15 individuals who underwent reduction mammoplasty surgeries and had no personal or familial history of cancer (<u>R</u>eduction <u>M</u>ammoplasty cohort, RM). Written informed consent was obtained from all enrolled individuals. The study was approved by the the Bioethical Committee at the Collegium Medicum, Nicolaus Copernicus University in Toruń (approval number KB509/2010) and by the Independent Bioethics Committee for Research at the Medical University of Gdańsk (approval number NKBBN/564/2018 with multiple amendments), recruited and enrolled all donors under informed and written consent, collected, and stored all tissue samples. A significant age difference was observed between BCAP, BCUS, and RM cohorts (Kruskal-Wallis test, p=7.7e-05), with the BCAP and BCUS cohorts being significantly older than the control group (Kruskal–Wallis H test, p=0.000034 and p=0.00036 for BCAP and BCUS respectively). However, no significant difference was observed between the BCAP (median age: 62, range: 23-85) and BCUS median age: 65, range: 37-84) cohorts (Kruskal–Wallis H test, p=0.082). A total of 415 samples, including PT, UM, UMD, BL, or SK from all three cohorts were collected by the Oncology Centre in Bydgoszcz, Jagiellonian University Hospital in Cracow, and the University Clinical Centre in Gdańsk, with the necessary ethical approvals and written informed consent from participants and deposited in the biobank of our unit at the Medical University of Gdańsk, along with clinical data, including follow-up information (Table 1, Supplementary Table 1). Distal UM samples (UMD, 1.5-3 cm from PT, median 2.35 cm), available for 7 BCAP patients, were collected and included in the downstream confirmatory analysis, however, they were not initially sequenced. For the RM cohort, sets of UM and BL samples were included. UM samples were located at least 1 cm away from the corresponding PT. All collected samples were frozen at -80°C. Detailed tissue-collecting protocols were previously described by Filipowicz et al^17^. All fragments prepared for molecular analysis were histologically evaluated by expert pathologists to identify tumor fragments (PT) and confirm the normal histology of UM and SK samples. DNA isolation from tissue lysates and whole blood was performed as previously described^17^.

**Table 1.** Summarized clinicopathological features of breast cancer patients included in the Breast Cancer Adverse Prognoses (BCAP) and the Breast Cancer Un-Selected (BCUS) cohorts. Matched and primary tumor (PT) and uninvolved mammary gland (UM, ≥1 cm) samples were collected from two breast cancer cohorts, i.e. 77 individuals characterized with adverse outcomes (BCAP cohort) and 49 individuals recruited without any pre-selection criteria related to prognosis (BCUS cohort). Whole peripheral blood (BL) or skin (SK) samples (if BL was not available) were collected as reference samples to distinguish between post-zygotic and germline variants. Distal UM samples (UMD, 1.5-3 cm from PT, median 2.35 cm), available for 7 BCAP patients, were included. The detailed sampling design is described in Materials and Methods. An overview is also available in Figure 1. *Death status refers to patients who succumbed to the disease (patient with ID BCAP61 died from non-oncological reasons). Detailed clinicopathological information for BCAP and BCUS cohorts is provided in Supplementary Table 1.

|  | **BCAP**<br>**cohort** | **BCUS cohort** |
| --- | --- | --- |
| **Number of individuals** | 77 | 49 |
| **Age (median, range)** | 62, 23-85 | 65, 37-84 |
|  |  | p value = 0.082 |
| **Collected samples** | 238 | 147 |
| Primary Tumor, PT | 77 | 49 |
| Uninvolved mammary gland, UM | 77 | 49 |
| Distal fragment of uninvolved mammary gland,<br>UMD | 7 | - |
| Reference sample<br>(whole peripheral blood, BL or skin, SK) | 77 | 49 |
| **Histology** |  |  |
| Invasive ductal carcinoma, IDC | 59 | 40 |
| Invasive lobular carcinoma, ILC | 3 | 4 |
| IDC - ILC | 6 | 1 |
| other | 9 | 4 |
| **Receptors** |  |  |
| Estrogen, ER (positive / negative / not available) | 57 / 20 | 43 / 5 / 1 |
| Progesterone, PR (positive / negative / not<br>available) | 43 / 34 | 44 / 4 / 1 |
| HER2 (positive / negative / not available) | 16 / 56 / 5 | 5 / 43 / 1 |
| **Subtype** |  |  |
| Luminal A | 14 | 22 |
| Luminal B | 37 | 21 |
| HER-2 enriched | 9 | 2 |
| Triple-negative breast cancer, TNBC | 11 | 1 |
| Not available | 6 | 3 |
| **Follow-up information** |  |  |
| Recurrence (yes / no) | 50 / 27 | 5 / 44 |
| Second cancer (yes / no) | 26 / 51 | 0 / 49 |
| Death* (yes / no) | 45 / 31 | 3 / 46 |

### Whole exome sequencing, data analysis, variant detection, and validation with independent methods

WES analyses were performed using the Agilent SureSelectXT Human All Exon V7 capture kit for sequencing library construction followed by 150 bp paired-end sequencing on the HiSeq Illumina platform (Illumina, San Diego, CA), outsourced to Macrogen Europe (Amsterdam, The Netherlands). Sequencing coverage was 200x on average, with at least 100x on target. The sequencing coverage and quality statistics for each sample are summarized in Supplementary Table 2A.

FASTQ files were inspected and processed with *Trim Galore!* (v0.6.7) (https://www.bioinformatics.babraham.ac.uk/projects/trim_galore/) to remove Illumina-specific adapter sequences and poor-quality reads when necessary. After converting FASTQ files to BAM format and extracting read groups from the raw data, reads were processed using GATK4 Best Practices (v4.0) (https://github.com/gatk-workflows/seq-format-conversion,https://github.com/gatk-workflows/gatk4-data-processing). The reads were mapped to the human genome (hg38) using the BWA-MEM tool (http://bio-bwa.sourceforge.net). Octopus (v0.7.4), in cancer mode, was used for variant calling. The cancer calling model can jointly genotype multiple samples from the same individual, using a reference sample (whole peripheral blood or skin when blood was not available) to distinguish between post- zygotic and germline variants (https://luntergroup.github.io/octopus/). Framesift insertions/deletions, nonsense, and missense variants located in exons were included in the analysis. A random forest filtering approach was implemented to minimize false calls. Variants in reads with poor mapping quality (<30), and variants supported by high-quality bases (≥30) in fewer than five reads were excluded from the analysis. Variants were annotated using ANNOVAR (https://annovar.openbioinformatics.org/) (last updated on 07.07.2020 and accessed between 06.2022-08.2022) and wANNOVAR (https://wannovar.wglab.org/) (last accessed on 21.05.2024), using the MANE SELECT transcript for investigated genes (https://www.ensembl.org/info/genome/genebuild/mane.html). A brief overview of filtering strategies implemented for identifying post-zygotic and germline variants related to breast cancer, within BCAP, BCUS, and RM cohorts and validation experiments for selected post-zygotic variants of BCAP patients is available in Supplementary Figure 2.

### Post-zygotic variants

Only variants with sequencing depth ≥ 50 and tissue allele frequency ≥ 0.03 were included in the analysis. Variants were filtered based on their annotation in ClinVar and InterVar databases; variants reported as “pathogenic”, “likely pathogenic”, “uncertain significance”, or “conflicting interpretations of pathogenicity” were included. In parallel, variants described in the COSMIC database (Cosmic_95coding) were incorporated. Missense variants documented in the COSMIC database, but annotated as “benign” or “likely benign” in ClinVar or InterVar databases, were excluded. Variants in genomic regions with known read-through transcription between adjacent genes, or those spanning multiple genes (e.g., *P2RY11; PPAN-P2RY11* and *KIR2DL1; KIR2DS5; LOC112267881*), were also excluded.

The remaining variants were filtered by their frequency in the general population, retaining only those with a minor allele frequency (MAF) ≤ 0.001 across all gnomAD populations (“popmax”) or not listed in gnomAD (v2.1.1) (Supplementary Table 3). Variants were further categorized as truncating (Supplementary Table 4) or missense (Supplementary Table 5). Truncating variants were considered pathogenic, regardless of their annotation in ClinVar or InterVar. For missense variants, *in-silico* analyses using the REVEL tool^18^ (with a threshold score of 0.75) were performed. In summary, truncating variants, variants classified as “pathogenic” or “likely pathogenic” in ClinVar, and missense variants with conflicting interpretations or annotated as “pathogenic” in ClinVar without assertion criteria, but having a REVEL score ≥ 0.75 were deemed pathogenic.

Variants affecting breast cancer-related genes were prioritized for further investigation based primarily on their ClinVar annotation. Variants annotated as “pathogenic” or “likely pathogenic” in ClinVar and missense variants reported in ClinVar as variants of uncertain significance or with conflicting interpretations (or without assertion criteria provided), with a REVEL score of ≥ 0.75 were included. Truncating variants affecting known tumor suppressor genes (i.e. *KMT2C*^19^, *TBX3*^20^, and *TP53*^21^) were also incorporated even if absent in ClinVar (Supplementary Table 6). UM, UMD, PT, and SK samples of 16 BCAP patients were subjected to further investigation (Sanger sequencing or High-Resolution Melting, Duplex sequencing) to verify or explore the presence of selected variants (Table 2, Supplementary Tables 7 and 8, Supplementary Figure 3).

**Table 2.** Pathogenic post-zygotic variants within the uninvolved mammary gland (UM) samples of breast cancer patients with adverse prognoses (BCAP cohort), selected for further investigation/validation. Presented variants, identified via Whole Exome sequencing, were corroborated with either Sanger sequencing/High-Resolution Melting, or Duplex sequencing. ^a^Variant annotation provided for the basic isoform of the transcript. ^b^Pathogenicity classification according to the ClinVar database. ^c^ID of the variant in the COSMIC (Cosmic_95 coding) database. ^d^rsIDs in dbSNP build 150. ^e^Individual ID and Variant Allele Frequency (VAF) for UM samples. Detailed description of selected post-zygotic variants is provided in Supplementary Table 6. Confirmation of post-zygotic variants by Sanger sequencing/High-Resolution Melting, or Duplex sequencing is provided in Supplementary Tables 7 and 8, respectively, and Supplementary Figure 3. SS – Sanger sequencing. HRM – High-Resolution Melting. DS – Duplex sequencing. n.a.- not available. *variants were also detected in the distal uninvolved mammary gland sample (UMD) of selected patients.

| Gene | Variant<sup>a</sup> | ClinVar<sup>b</sup> | COSMIC ID<sup>c</sup> | AVSNP150<sup>d</sup> | Individual ID and UM sample VAF | Confirmation |
| --- | --- | --- | --- | --- | --- | --- |
| *AKT1* | c.49G>A (p.Glu17Lys) | Pathogenic | ID=COSV62571334 | rs121434592 | BCAP32 (0.6%), BCAP66* (0.7%) | SS / HRM |
| *PIK3CA* | c.1624G>A (p.Glu542Lys) | Pathogenic | ID=COSV55873227 | rs121913273 | BCAP56 (0.7%), BCAP45 (12%) | SS / HRM |
| *PIK3CA* | c.3140A>G (p.His1047Arg) | Pathogenic | ID=COSV55873195 | rs121913279 | BCAP15 (0.08%), BCAP31* (19%), BCAP36 (0.3%), BCAP53* (0.7%) | SS / HRM / DS |
| *PIK3CA* | c.3140A>T (p.His1047Leu) | Pathogenic | ID=COSV55873401 | rs121913279 | BCAP54* (0.5%) | DS |
| *PTEN* | c.388C>T (p.Arg130*) | Pathogenic | ID=COSV64288463 | rs121909224 | BCAP15 (0.7%) | SS / HRM |
| *TBX3* | c.371_372insTGGT (p.Ile125Profs*14) | n.a. | ID=COSV57471668 | n.a. | BCAP44 (12%) | SS / HRM |
| *TP53* | c.151G>T (p.Glu51*) | Pathogenic | ID=COSV52694020 | n.a. | BCAP58* (16%) | SS / HRM |
| *TP53* | c.227del (p.Ala76Aspfs*47) | n.a. | ID=COSV52728465 | n.a. | BCAP54 (0.5%) | DS |
| *TP53* | c.329G>C (p.Arg110His) | Pathogenic | ID=COSV52668419 | rs11540654 | BCAP45 (0.8%) | DS |
| *TP53* | c.637C>T (p.Arg213*) | Pathogenic | ID=COSV52665560 | rs397516436 | BCAP01* (0.8%), BCAP48* (0.6%) | DS |
| *TP53* | c.711G>A (p.Met237Ile) | Pathogenic | ID=COSV52661887 | rs587782664 | BCAP15 (0.8%) | SS / HRM |
| *TP53* | c.1024C>T (p.Arg342*) | Pathogenic | ID=COSV52665487 | rs730882029 | BCAP38 (0.5%), BCAP47 (0.7%) | DS |
| *TP53* | c.1025G>C (p.Arg342Pro) | Pathogenic/<br>Likely pathogenic | ID=COSV52690857 | rs375338359 | BCAP57 (0.6%) | DS |

### Germline variants

For germline variant detection, only variants in high- and moderate-penetrance breast cancer susceptibility genes were included in the study, as defined by the NCCN Clinical Practice Guidelines in Oncology^22^ (Version 1.2023, September 7, 2022). The list of genes of interest includes: *ATM* (MIM *607585)*, BRCA1* (MIM *113705)*, BRCA2* (MIM *600185)*, BARD1* (MIM *601593)*, BRIP1* (MIM *605882)*, CHEK2* (MIM *604373)*, CDH1* (MIM *192090)*, PALB2* (MIM *610355)*, PTEN* (MIM *601728)*, TP53* (MIM *191170)*, NF1* (MIM *613113)*, STK11* (MIM *602216)*, RAD50* (MIM *604040)*, RAD51C* (MIM *602774)*, RAD51D* (MIM *602954) and additionally *PIK3CA* (MIM *171834). Variants were filtered based on their frequency in the general population: variants with minor allele frequency (MAF) ≤ 0.01 across all gnomAD populations (“popmax”) or not noted in the database (gnomAD v2.1.1) were included. Evidence according to the American College of Medical Genetics and Genomics and the Association for Molecular Pathology recommendations^23^ was included to describe all germline pathogenic variants. Specifically, the evaluation of identified *BRCA1* and *BRCA2* variants was performed according to the Evidence-based Network for the Interpretation of Germline Mutation Alleles (ENIGMA) *BRCA1* and *BRCA2* Variant Curation Expert Panel^24^ (Version 1.1.0) (Clinical Genome Resource, https://www.clinicalgenome.org/affiliation/50087/, https://cspec.genome.network/cspec/ui/svi/doc/GN092, https://cspec.genome.network/cspec/ui/svi/doc/GN097). Pathogenic germline variants meeting the study’s criteria, identified within breast cancer patients of the BCAP and BCUS cohorts are described in Supplementary Table 9.

### Duplex sequencing

UM samples from 11 BCAP patients were selected to investigate the presence of low-frequency *PIK3CA* and *TP53* variants beyond the detection limits of Sanger sequencing and High-Resolution Melting and a single, higher-frequency *TP53* variant, i.e. c.151G>T (p.Glu51*), located in a difficult GC-rich region. (Table 2, Supplementary Table 8). Additionally, UMD samples, available for 6 of those patients, were included to explore further the distribution of selected variants in a more distant seemingly normal mammary tissue. Duplex sequencing was performed as previously described^15,25^.

### Duplex sequencing data analysis

Raw duplex sequencing data were analyzed using the Snakemake-based Duplex-seq-Pipeline (v1.1.4) (https://github.com/Kennedy-Lab-UW/Duplex-Seq-Pipeline) as previously described^26^. The sequencing coverage and quality statistics for each sample are summarized in Supplementary Table 2B.

### Statistical analysis

All statistical analyses were carried out with in-house developed scripts using R studio version 4.1.2 (2021-11-01). Packages *pheatmap* (version 1.0.12) and *ggplot2* (version 3.4.1) were used for plotting. Statistical significance of differences between two or multiple groups was tested using the Mann– Whitney U test or the Kruskal–Wallis H test respectively. Statistical significance of features between multiple groups was tested with the Hypergeometric test or Fisher’s exact test. Hazard Ratios were calculated using the *coxph* function from package *survival* (version 3.5-5). Kaplan-Meier analysis was performed using the *survfit* and *ggsurvplot* functions from *survminer* package (version 0.4.9) and groups were tested with log-rank test. Differences were considered significant at p < 0.05.

## RESULTS

### Truncating post-zygotic variants in dosage-sensitive genes predominate in BCAP compared to BCUS and RM cohorts

We examined sets of UM and PT samples from all breast cancer patients to identify variants related to breast cancer (Figure 1a). Whole peripheral blood (BL) or skin (SK), if blood was not available, served as a reference sample, and was used to distinguish between post-zygotic and germline variants (Figure 1b). The details of post-zygotic variants in BCAP, BCUS, and RM cohorts meeting the study’s cut-off criteria (Materials & Methods and Supplementary Figure 2) are summarized in Supplementary Table 3. Briefly, all truncating variants were included for manual curation, while non-truncating variants were filtered based on their annotation within ClinVar or InterVar databases as “pathogenic”, “likely pathogenic”, “uncertain significance”, or “conflicting interpretations of pathogenicity”, or their presence in the COSMIC database (cosmic_95coding). Additionally, variants were filtered by their frequency in the general population including only those with a minor allele frequency (MAF) ≤ 0.001 across all gnomAD populations (“popmax”) or not noted in the database (gnomAD v2.1.1).

**Figure 1.**
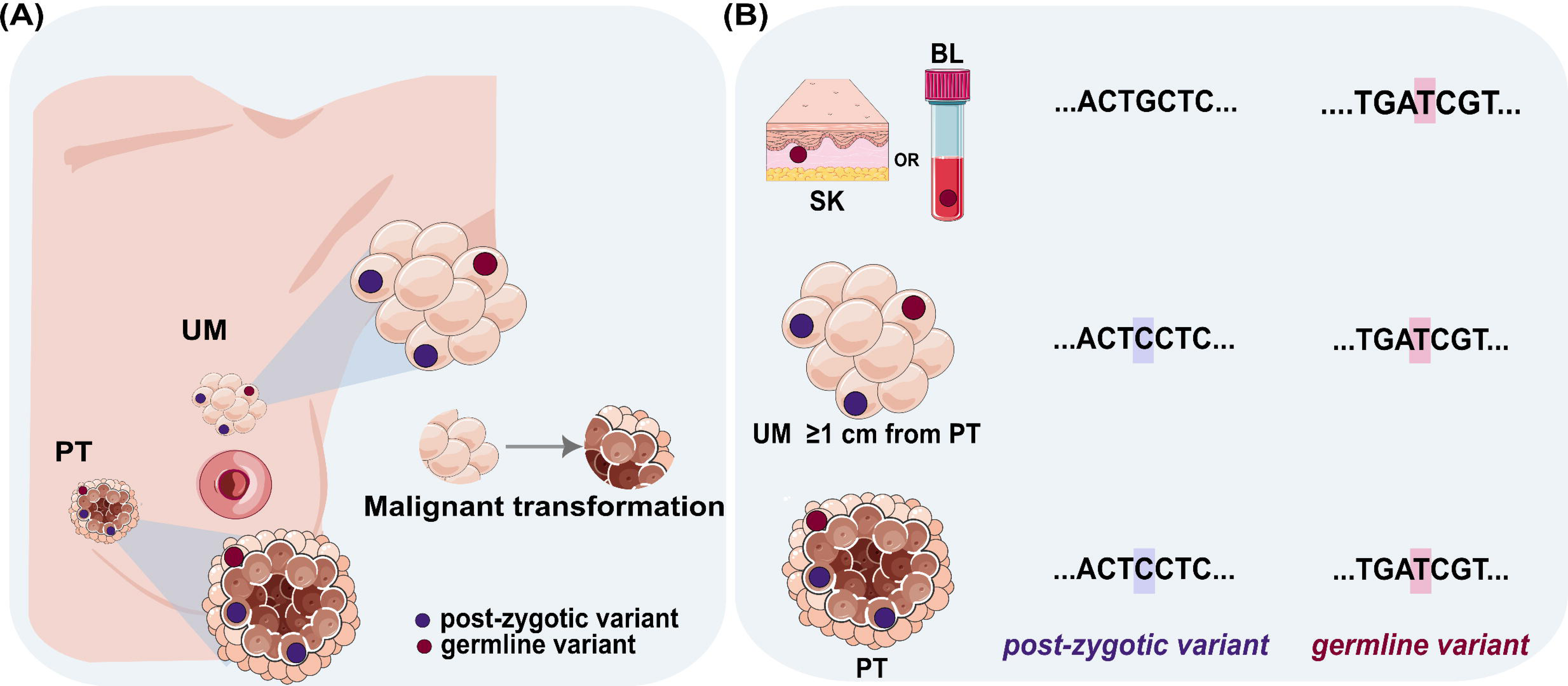
Graphical representation of the sampling design used for variant investigation. (**A) Sampling design.** Matched fresh-frozen uninvolved mammary gland (UM) and primary tumor (PT) samples were collected from all breast cancer patients to identify variants according to study criteria (Materials & Methods). UM samples were located at least 1 cm away from the corresponding PT. Control mammary gland samples were obtained from individuals who underwent reduction mammoplasty surgeries and had no history of cancer (Reduction Mammoplasty cohort, RM). (**B) Investigation of post- zygotic and germline variants.** Matched peripheral blood (BL) or skin (SK) samples were collected for each individual as reference samples to distinguish between post-zygotic and germline variants. Post- zygotic variants were identified as those that were absent from the reference samples.

Within the BCAP cohort, 167 distinct variants were identified in UM samples of 41 patients. The corresponding numbers for the BCUS were strikingly lower, i.e. 56 variants were identified in the UM samples of 24 patients. The RM cohort presented 10 distinct variants in seven unrelated individuals, but all were either variants of uncertain significance or not reported in the ClinVar/InterVar databases. Variants were further categorized based on their impact on the encoded protein i.e. truncating and non-truncating. Notably, truncating nonsense (n=25) and frameshift (n=12) variants, were exclusive to the BCAP cohort (Supplementary Table 4). These alterations were responsible for transcript elimination via nonsense-mediated mRNA decay^27^ and were deemed pathogenic, irrespective of their annotation in the ClinVar and InterVar databases. For missense variants in BCAP, BCUS, and RM UM samples, further analyses were performed using multiple *in-silico* predictors (Supplementary Table 5).

In the BCAP cohort, 29% (49/167) of the identified variants were deemed pathogenic. This list included truncating variants (n=37), missense variants annotated as pathogenic/likely pathogenic (n=8), and missense variants reported as uncertain significance or pathogenic/likely pathogenic in ClinVar, lacking assertion criteria but showing evidence of pathogenicity according to *in-silico* analyses (n=4) (Supplementary Tables 4 and 5). *In-silico* analyses for missense variants were conducted using the REVEL tool^18^, with a threshold set to 0.75 (accessed between June 2022 and August 2022 via ANNOVAR software). Notably, nearly one-quarter (24%, 12/49) of the pathogenic BCAP variants were detected only in the UM samples and were absent in the corresponding PTs. In comparison, the BCUS cohort had seven pathogenic variants, representing 13% of the total identified variants (n=56). These included variants reported as pathogenic (n=4), and missense variants classified as uncertain significance or pathogenic/likely pathogenic in ClinVar without assertion criteria, showing evidence of pathogenicity according to REVEL (n=3) (Supplementary Table 5). Nearly half (43%, 3/7) of the pathogenic BCUS variants were identified only in UM samples. The UM samples from the BCAP cohort were significantly enriched for pathogenic post-zygotic variants (a combination of truncating variants and those non-truncating variants, classified as pathogenic by ClinVar/REVEL) compared to those from the BCUS cohort (Hypergeometric test, p=0.0008578).

To select post-zygotic variants potentially linked to breast cancer, we focused on pathogenic variants as defined by the ClinVar database. Variants annotated as pathogenic or likely pathogenic in ClinVar, reviewed by expert panels, or submitted by multiple parties without discordance are listed in Supplementary Table 6 for both BCAP and BCUS cohorts. Missense variants classified as uncertain significance, pathogenic/likely pathogenic without assertion criteria, or with conflicting interpretations in ClinVar with a REVEL score ≥ 0.75 were also included. Truncating variants in known tumor-suppressor genes implicated in breast cancer, such as *KMT2C*^28^, *TBX3*^20^, and *TP53*^21^ were included in the same table even if they were absent from the ClinVar database (Supplementary Table 6).

We identified several variants affecting dosage-sensitive genes. These included deleterious variants in the tumor suppressors *KMT2C*^28^, *PTEN*^29^, *PTCH1*^30^, *TBX3*^20^, and *TP53*^21^ as well as activating variants in oncogenes *AKT1*^31^ and *PIK3CA*^32^ identified in BCAP UM samples (Supplementary Table 6). Oncogenes such as *SF3B1*^33^, *HRAS*^34^, and *GNAS*^35^, and genes with a dual role in cancer (*RUNX1*^36^) were affected solely in the BCUS cohort, with only the latter two being dosage-sensitive (Supplementary Table 6). *PIK3CA* was the only gene recurrently affected in both BCAP and BCUS cohorts.

### Recurrence, coexistence, spatial distribution in the breast, and validation of variants

In the BCAP cohort, pathogenic variants in two driver genes, *PIK3CA* and *TP53*, were predominant across all subtypes of invasive cancer. *PIK3CA,* which encodes the catalytically active p100alpha isoform, is a key regulator of cell proliferation and growth receptor signaling cascades^37^. We detected three distinct pathogenic post-zygotic *PIK3CA* variants in UM samples: c.1624G>A (p.Glu542Lys), c.3140A>G (p.His1047Arg), and c.3140A>T (p.His1047Leu), found in two, four and one unrelated individuals, respectively (Supplementary Table 5). Another *PIK3CA* variant, c.3012G>T (p.Met1004Ile), was found in the UM of a single BCAP cohort individual. However, this variant reported only once as of uncertain significance in the ClinVar database and with a REVEL score of 0.437 suggesting it might be benign, was classified as a variant of uncertain significance due to limited evidence (Supplementary Table 5). *PIK3CA* c.3140A>G (p.His1047Arg) and c.3140A>T (p.His1047Leu) variants, located at the commonly mutated *PIK3CA* site in breast cancer, were also observed in UMs of the BCUS cohort (Figure 2, Supplementary Table 5).

**Figure 2.**
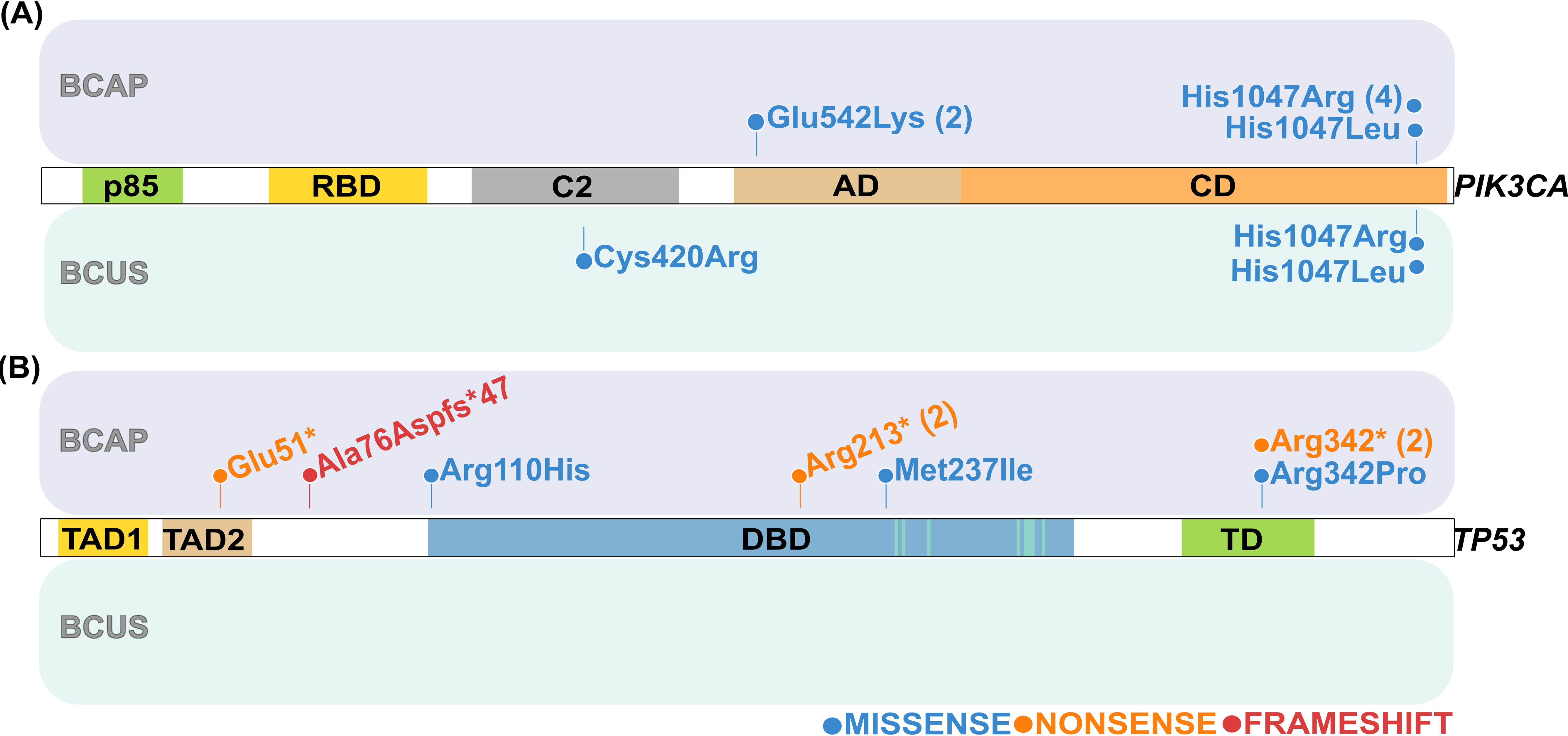
Post-zygotic (A) *PIK3CA* and (B) *TP53* variants detected in the uninvolved mammary gland (UM) and primary tumor (PT) samples of breast cancer patients with adverse prognoses and patients recruited without any prognoses bias (BCAP and BCUS cohorts, respectively). Lollipop plots represent post-zygotic variants of *PIK3CA* and *TP53* genes detected by Whole Exome Sequencing (WES). The upper panel represents variants detected in UM samples of BCAP patients. The lower panel represents variants detected in UM samples of BCUS patients. *TP53* variants were found exclusively in the BCAP cohort. All identified variants have been reported in the COSMIC database (https://cancer.sanger.ac.uk/cosmic). Detailed descriptions of variants can be found in Supplementary Table 6. p85 - p85 binding domain, RBD - Ras-binding domain, C2 - C2 domain, AD - accessory domain, CD - catalytic domain. TAD1, TAD2 - transcription activation domain 1 and 2, DBD - DNA-binding domain, DNA-binding sites are marked with green lines, TD - tetramerization domain. Lollipop plots were prepared based on the images generated with the Protein paint application^53^. () indicates the number of patients in whom the variant was identified.

*TP53* is the most commonly mutated gene in various human cancers and its normal protein function is frequently compromised in many types of malignancies^38^. We detected seven *TP53* variants in the normal mammary gland samples of nine BCAP patients, including two recurrent variants: six pathogenic or likely pathogenic (c.151G>T [p.Glu51*], c.329G>C [p.Arg110His], c.637C>T [p.Arg213*], c.711G>A [p.Met237Ile], c.1024C>T [p.Arg342*], and c.1025G>C [p.Arg342Pro]), and one frameshift variant c.227del (p.Ala76Aspfs*47), not previously reported in the ClinVar database (Supplementary Table 6). Importantly, no *TP53* variants were observed in the normal mammary gland samples of the BCUS cohort (Figure 2). Pathogenic variants in *AKT1*, *PIK3CA*, *PTEN*, *TBX3*, and *TP53* in 16 BCAP patients were selected for validation with independent methods (Table 2, Supplementary Figure 2). The presence of *PIK3CA* variants c.3140A>G (p.His1047Arg) (in three patients), c.1624G>A (p.Glu542Lys) (in two patients) and *TP53* c.711G>A (p.Met237Ile) (in a single patient) was confirmed by Sanger sequencing or High-Resolution Melting in the UM samples of six BCAP patients (Table 2, Supplementary Table 7, Supplementary Figure 3). Notably, the *PIK3CA* c.3140A>G (p.His1047Arg) and *TP53* c.711G>A (p.Met237Ile) variants co-existed in the UM sample of patient BCAP15. Additionally, two pathogenic variants, c.49G>A (p.Glu17Lys) in *AKT1* and c.388C>T (p.Arg130*) in *PTEN*, were identified in the UM samples of three BCAP patients and subsequently confirmed via Sanger sequencing or High-Resolution Melting (Table 2, Supplementary Table 7, Supplementary Figure 3). The presence of the *AKT1* variant was confirmed in the UM samples of two patients and the UMD of one of them, indicating a broad spatial distribution of this variant in that particular patient. Finally, the *TBX3* c.371_372insTGGT (p.Ile125Profs*14) variant was confirmed in the UM sample of a single patient (Table 2, Supplementary Table 7, Supplementary Figure 3). We further employed duplex sequencing to validate the presence of extremely low-frequency *PIK3CA* and *TP53* variants and examine their tissue/spatial distribution. We selected 11 individuals: BCAP01, BCAP31, BCAP36, BCAP38, BCAP45, BCAP47, BCAP48, BCAP53, BCAP54, BCAP57, and BCAP58 based on the presence of *PIK3CA* and *TP53* variants in proximal UM samples from WES data. UMD samples available for 6 BCAP patients, located at a greater distance from the PTs and not included in the original WES run, were also analyzed (Table 2). Ultra-deep targeted duplex sequencing with a mean coverage of 4789x confirmed the following low-frequency (low as 1.34%) pathogenic variants in *PIK3CA*: c.3140A>G (p.His1047Arg) and c.3140A>T (p.His1047Leu) in the UM samples from two patients, and revealed the presence of c.3140A>G (p. His1047Arg) variant in the UMD samples of another two patients (Table 2, Supplementary Table 8). Furthermore, duplex sequencing confirmed the presence of six *TP53* variants (c.151G>T [p.Glu51*], c.227del [p.Ala76Aspfs*47], c.329G>C [p.Arg110His], c.637C>T [p.Arg213*], c.1024C>T [p.Arg342*], and c.1025G>C [p.Arg342Pro]) in UM tissues of eight patients, and revealed the presence of c.151G>T (p.Glu51*), c.227del (p.Ala76Aspfs*47), and c.637C>T (p.Arg213*) variants in the paired UMD tissues of four patients (Table 2, Supplementary Table 8). An overview of validated variants in *AKT1*, *PIK3CA*, *PTEN*, *TBX3,* and *TP53* genes along with follow-up information for the corresponding patients is provided in Figure 3.

**Figure 3.**
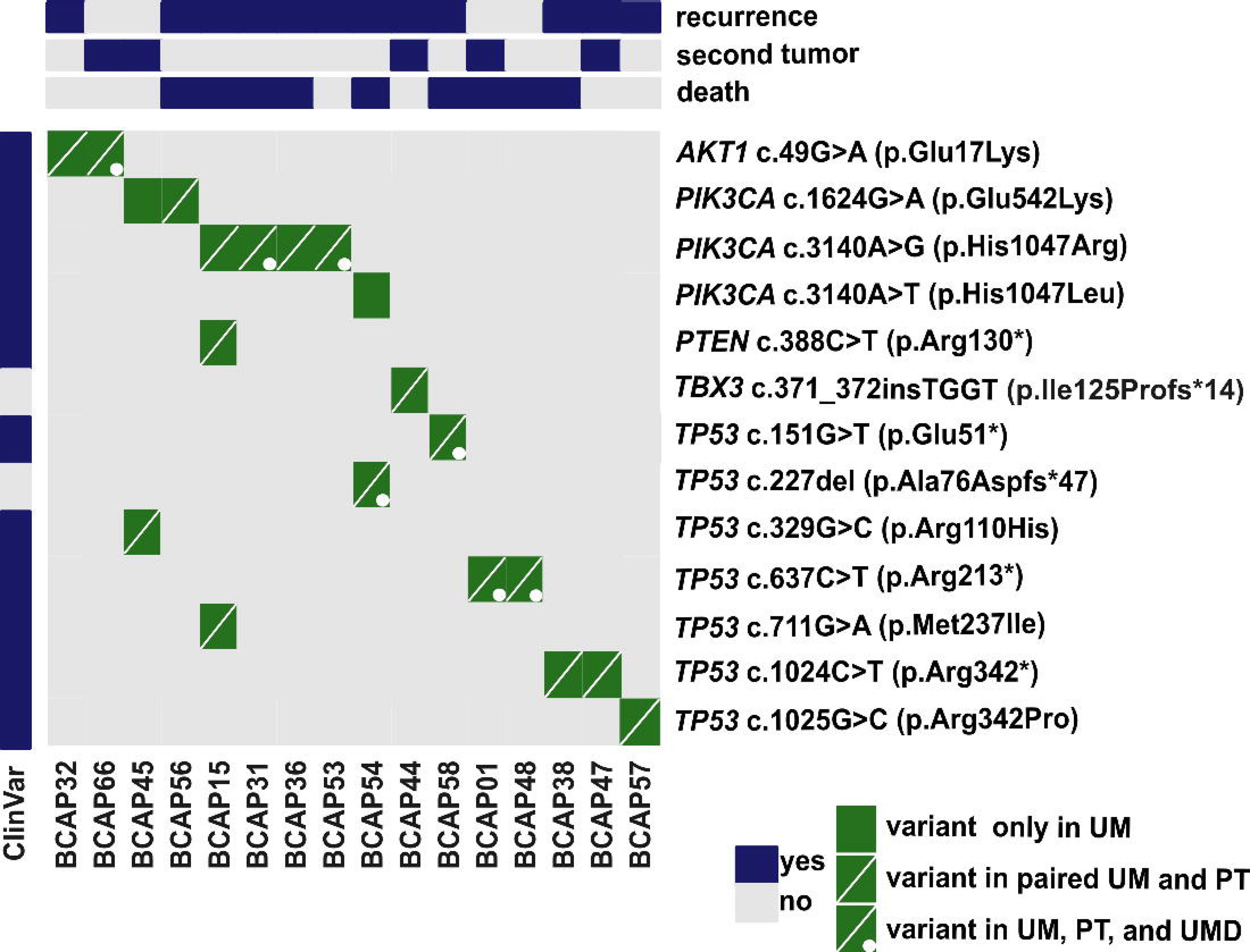
Pathogenic post-zygotic variants in breast cancer patients with adverse prognoses. Variants were confirmed via Sanger sequencing/High-Resolution Melting or Duplex sequencing (Materials and Methods, Table 2, Supplementary Tables 7 and 8, Supplementary Figure 3). Variant presence in the ClinVar database and follow-up information for the corresponding patients are included. Detailed clinicopathological information for presented patients is provided in Supplementary Table 1. A full description of detected variants is provided in Supplementary Table 6. PT – primary tumor. UMD – uninvolved mammary gland at a further distance from corresponding PT.

### Spectrum of germline pathogenic variants in the two breast cancer cohorts

All breast cancer cases included in our study were reported as sporadic based on the family history of the patient, however, genetic testing results were not available prior to recruitment. We analyzed BL or SK samples from each participant to screen for pathogenic or likely pathogenic germline variants across all cohorts (Materials & Methods, Supplementary Figure 2). In the BCAP cohort, 14 of 77 individuals (18%) carried germline pathogenic variants in known breast cancer-associated genes^22^. These included c.4186C>T (p.Gln1396*), c.4689C>G (p.Tyr1563*), c.5179A>T (p.Lys1727*), and c.5266dup (p.Gln1756Profs*74) in *BRCA1*, c.5645C>A (p.Ser1882*), c.6591_6592del (p.Glu2198Asnfs*4), and c.9382C>T (p.Arg3128*) in *BRCA2*, c.172_175del (p.Gln60Argfs*7) and c.1671_1674del (p.Ile558Lysfs*2) in *PALB2*, and c.3233_3236del (p.Lys1079Valfs*28) in *RAD50*. Only *BRCA1* c.5266dup (p.Gln1756Profs*74) and *PALB2* c.172_175del (p.Gln60Argfs*7) were recurrent, observed in four and two unrelated individuals, respectively. This incidence surpasses the rates from other studies where up to approximately 10% of reportedly sporadic cases turn out hereditary after molecular testing and likely reflects the aggressive outcomes in these cases^15,21,39^. Four individuals (4/14, 29%) with germline pathogenic variants also carried pathogenic post-zygotic variants in known, curated cancer- related genes^19^ in their UM samples. The germline variants in these cases were found in *BRCA1* (four cases) and *RAD50* (one case). The corresponding post-zygotic variants were identified in *PIK3CA* or *TP53*. *BRCA1* c.5266dup (p.Gln1756Profs*74) was the only variant observed in a single patient from the BCUS cohort. In the control group, no individuals were found to carry germline pathogenic or likely pathogenic variants in genes associated with breast cancer. Pathogenic germline variants in high- and moderate-penetrance breast cancer susceptibility genes identified in the BCAP and BCUS cohorts are detailed in Supplementary Table 9.

### Pathogenic post-zygotic variants in patients with recurrent disease affect the survival rate

We used Kaplan-Meier plots to evaluate survival probabilities and compare patients with recurrence (n=53) to those without recurrence (n=72) in both the BCAP and BCUS cohorts. Overall, patients with recurrence had significantly lower survival probabilities (log-rank test, p=0.00017) (Supplementary Figure 4A), with a hazard ratio of 2.44 (95% CI: 1.07-5.54, p=0.0337), indicating more than twice the risk of death compared to the non-recurrence group.

Due to the shorter follow-up period for the BCUS cohort (2 years) compared to the BCAP cohort (10 years), we focused on the first 24 months post-diagnosis. During this period, recurrence patients in both cohorts (n=53) had significantly lower survival probabilities compared to non-recurrence patients (n=71) (log-rank test, p<0.0001) (Supplementary Figure 4B), with a hazard ratio of 4.85 (95% CI: 1.4-16.25, p=0.0105), indicating more than four times the risk of death for the recurrence group. Within the first 24 months, BCAP patients experienced significantly more recurrence events than BCUS patients (Fisher’s exact test, p=0.005488). In the BCAP cohort, patients with recurrence (n=48) had lower survival probabilities throughout the follow-up period compared to those without recurrence (n=28) (log-rank test, p=0.015). This pattern was also observed in the first 24 months (n=48 vs. n=27) (log-rank test, p=0.0088) (Supplementary Figures 4C and 4D).

Additionally, we assessed the impact of pathogenic post-zygotic and germline variants on the survival of patients with recurrent disease. Figure 4 displays Kaplan-Meier curves based on the presence and type of pathogenic variants. Survival probabilities differ significantly across studied groups (log-rank test, p=0.024), indicating that the combination of pathogenic post-zygotic variants, pathogenic germline variants, and recurrence status significantly impact patient survival. Patients with pathogenic germline variants (green) had the shortest recurrence-free survival, with most recurrences occurring within the first 60 months. Notably, patients with pathogenic post-zygotic variants in breast cancer-specific genes (blue) also experienced recurrences, though less frequently and over a longer follow-up period, highlighting the impact of these variants on recurrence risk, albeit to a lesser extent than germline variants. Patients without pathogenic germline or postyzygotic variants (yellow) showed intermediate outcomes. In conclusion, while pathogenic germline variants have the most pronounced effect on survival rates, pathogenic post-zygotic variants seem to also play a notable role in influencing recurrence risk and patient outcomes, highlighting their importance in understanding disease progression.

**Figure 4.**
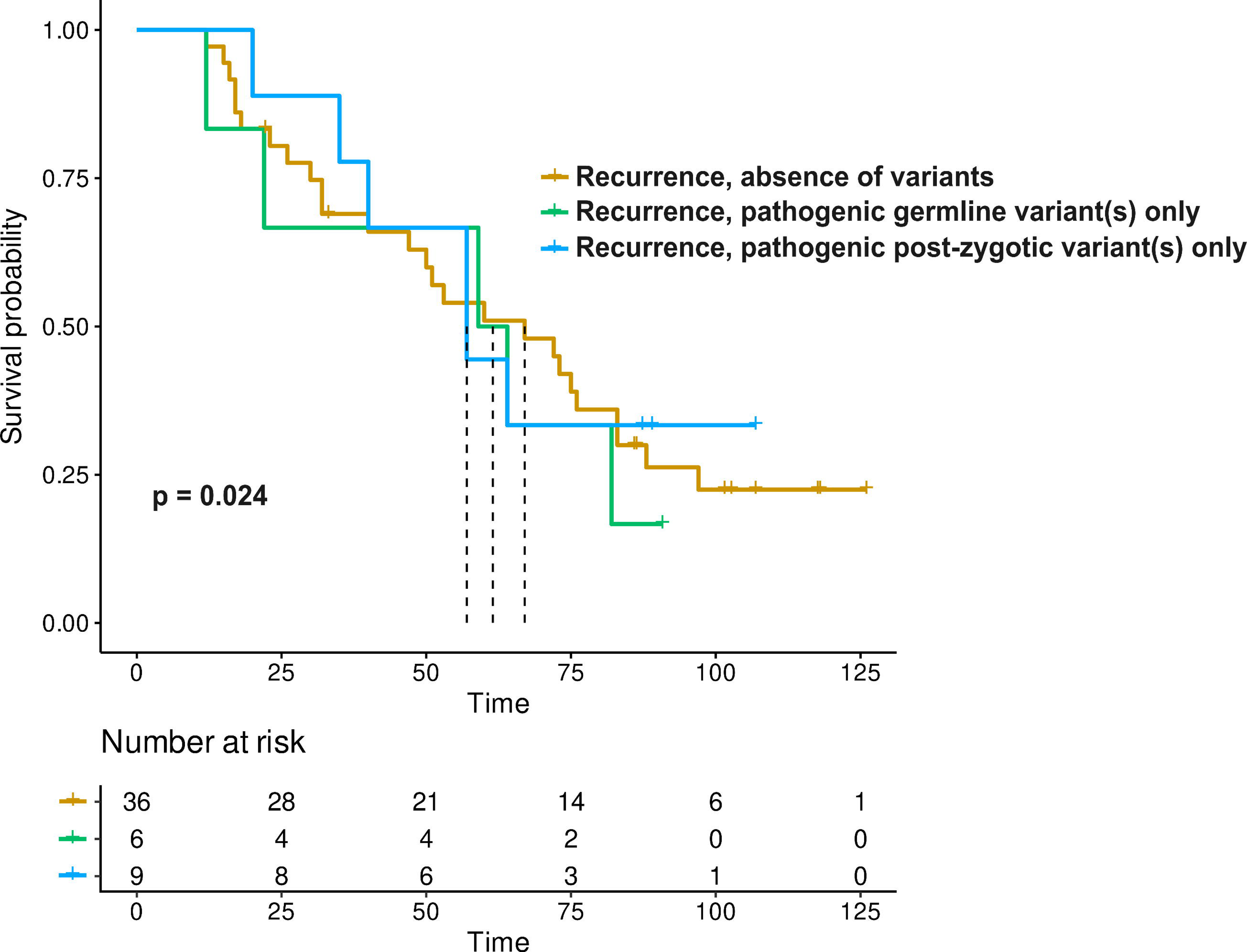
Kaplan-Meier survival curves of breast cancer patients with pathogenic variants and recurrent disease. The curves represent survival probabilities for different groups of patients from the BCAP cohort (breast cancer patients with adverse prognoses) and the BCUS cohort (breast cancer patients without specific prognosis criteria), stratified by the presence of recurrent disease and/or pathogenic germline or post-zygotic variants in breast cancer-specific genes. Survival time was measured from the date of diagnosis to death or the end of the follow-up period (10 years for BCAP and 2 years for BCUS). Detailed information on recurrence status and patient outcomes can be found in Supplementary Table 1. The x-axis represents time in months, and the y-axis represents the probability of survival. Vertical ticks on the curves indicate censored death events.

## DISCUSSION

The early detection and treatment of breast cancer, including its precursors, have shifted research focus from tumors to the normal mammary gland to deepen our understanding of the disease’s origins^40^. Genetic and transcriptomic studies have revealed a wide spectrum of alterations in critical breast cancer driver genes within the normal mammary gland of patients who have undergone BCS or mastectomy, compared to control tissues^11–15,41^.

Recognizing that histologically normal tissue may harbor early genetic changes, we screened for post- zygotic alterations in two similarly aged breast cancer cohorts (BCAP and BCUS, Kruskal–Wallis H test, p=0.082) with differing survival outcomes. We also included a significantly younger control group of individuals treated surgically for non-cancerous reasons (RM cohort, Kruskal–Wallis H test, p=0.000034 and p=0.00036 for BCAP and BCUS cohorts respectively).

Truncating variants (nonsense and frameshift) were found exclusively in the UM samples of patients with adverse prognoses (BCAP cohort) (Supplementary Table 4). In contrast, BCUS patients and RM controls showed only missense variants (Supplementary Table 5). UM samples from BCAP patients were significantly enriched for pathogenic post-zygotic variants (Hypergeometric test, p=0.0008578), affecting several known cancer-related genes genes^19^ i.e. *AKT1*, *KMT2C*, *PIK3CA*, *PTCH1*, *PTEN*, *TBX3*, and *TP53,* and almost a quarter were found exclusively in UM samples, absent from the corresponding PTs, suggesting early tumorigenic processes. These findings indicate that the presence of pathogenic post- zygotic alterations in UM samples could signal a higher risk for aggressive cancer, preceding clinical symptoms.

The high prevalence of *PIK3CA* and *TP53* variants in BCAP patients’ UM samples highlights their pivotal role in oncogenesis. The *PIK3CA* gene encodes the p110α catalytic subunit of phosphoinositide 3-kinase (PI3K), a key regulator of the PI3K/AKT signaling pathway, essential for cellular growth, proliferation, and survival and has a known oncogene function^37,39^. Missense variants in *PIK3CA*, especially in the accessory (p.Glu542Lys) and catalytic domains ([p.His1047Arg], [p.His1047Leu]), enhance kinase activity and promote oncogenic signaling. The recurrence of the p.His1047Arg variant, critical for PIK3CA function, underscores its significance in tumorigenesis. *PIK3CA* variants were common in both BCAP and BCUS cohorts, suggesting a fundamental role in breast cancer development. In contrast, *TP53* variants were found exclusively in the BCAP cohort, underscoring the gene’s crucial role in maintaining genomic integrity. *TP53* encodes the tumor protein p53, a key tumor suppressor involved in DNA repair, apoptosis, and cell cycle regulation^21^. Loss-of-function variants in *TP53* can inactivate its tumor-suppressing activity during oncogenesis^42–44^. The observed *TP53* variants included truncating variants (nonsense and frameshift) as well as missense alterations affecting the DNA-binding domain ([p.Arg110His], [p.Met237Ile]) and the tetramerization motif (p.Arg342Pro), highlighting the various ways p53 function can be disrupted, potentially leading to malignancy. The exclusive presence of *TP53* variants in the BCAP cohort may suggest a more aggressive disease phenotype and a worse prognosis associated with these variants.

*PIK3CA* and *TP53* variants co-occurred in three BCAP patients (BCAP15, BCAP45, BCAP54), suggesting a synergistic role in cancer progression. In particular, patient BCAP15 exhibited concurrent pathogenic variants in *PIK3CA* (p.His1047Arg), *TP53* (p.Met237Ile), and *PTEN* (p.Arg130*). *PTEN*, another critical tumor suppressor, negatively regulates the PI3K/AKT pathway^29^. The presence of alterations in all three genes within a single patient emphasizes the complex interplay of multiple oncogenic and tumor-suppressive pathways in breast cancer pathogenesis. This confluence of variants likely contributes to a more aggressive clinical course, underscoring the importance of comprehensive genetic profiling in understanding individual tumor biology.

While post-zygotic variants in *TP53* or *PIK3CA* have been observed in breast tumors, their consequences in normal mammary tissue are less clear. Some studies suggest a benign effect^45,46^. Healthy breast tissue accumulates alterations with age at an accelerating rate^47^ and is influenced by hormonal stimuli, undergoing cycles of expansion during puberty, pregnancy, and lactation^48^. Estrogen and its metabolites can cause DNA damage, increasing cellular stress and the risk of genetic alterations and cancer^48^. However, the accumulation of alterations alone does not cause cancer; observed tissue-specific patterns and the “ground state” theory suggest that quiescent stem cells with oncogenic variants rarely transform unless activated by developmental, aging, or injury factors, that normally resemble physiological mammary gland conditions^49^. The presence of such variants in ostensibly normal mammary gland tissue suggests they may represent early, pre-cancerous changes. Despite the mammary gland’s inherent multi- layer protection system against clonal expansions, surviving mutant clones can lead to large fields of mutated cells, i.e. field cancerization, thereby increasing cancer risk^50^.

A subset of the BCAP cohort (18%, n=14/77) and one patient from the BCUS cohort (2%, n=1/49) carried germline pathogenic variants in breast cancer genes (Supplementary Table 9). Among the BCAP patients with these variants, four had concurrent pathogenic post-zygotic variants in curated cancer genes, whereas 14 had only post-zygotic alterations in curated cancer genes and genes implicated in breast cancer (Supplementary Tables 6 and 9). Despite their differing genetic profiles, all BCAP patients experienced adverse outcomes within ten years of surgery highlighting the impact of these genetic variations on prognosis. The interaction between germline and post-zygotic variants remains unclear, as recent research indicates that the influence of germline variants on tumor behavior can vary significantly based on factors such as penetrance and lineage, with some variants exhibiting minimal or transient effects on tumor development^51^.

In the first 24 months post-diagnosis, the BCAP cohort had significantly more recurrence events than the BCUS cohort (Fisher’s exact test, p = 0.005488). Recurrence was associated with much lower survival probabilities across both cohorts. While pathogenic post-zygotic variants alone did not drastically alter survival rates, their impact became more severe when combined with disease recurrence, suggesting a link to disease aggressiveness. This highlights the importance of comprehensive genetic screening and vigilant monitoring of patients with pathogenic post-zygotic variants in disease-related genes to improve outcomes.

Current diagnostics primarily focus on identifying germline pathogenic variants in known breast cancer-associated genes to assess breast cancer risk and guide personalized therapy^22^. However, over 80% of breast tumors are not caused by inherited alterations^9^. Our study reveals that pathogenic post-zygotic variants, such as *PIK3CA* and *TP53* alterations, are often found in seemingly normal mammary gland tissue left behind after BCS, with allele frequencies ranging from 0.03 to 0.28. In some instances, these variants represent distinct clonal populations not present in the corresponding primary tumors (Supplementary Table 6), indicating the independent evolution of cell lineages within the mammary tissue and suggesting they are unlikely to be micrometastases. The timing of their emergence relative to the primary tumor - whether during early tumor progression or later—remains uncertain, but in both scenarios, they may contribute to recurrence in the breast or metastasis to other organs^52^. Understanding these dynamics is essential for improving diagnostic approaches and tailoring effective treatments.

Our study comes with certain limitations, particularly regarding the notable age differences between the breast cancer cohorts and individuals subjected to reduction mammoplasty surgeries. Recruiting age-matched control individuals poses a challenge, as those opting for cosmetic surgical treatments are typically younger. The incidence of breast cancer diagnosis among younger women is relatively infrequent with only about one out of eight invasive breast cancers being diagnosed in women under the age of 45^1^. Another limitation arises in recruiting healthy control individuals, given that approximately 13% of women are expected to develop invasive breast cancer during their lifetime and the precise onset of carcinogenesis remains unclear^1,2^. Here, control normal mammary glands were sampled from individuals without a personal or familial history of cancer undergoing plastic surgery. Hence, these samples represent the most appropriate, available control samples from a biological standpoint.

Our findings demonstrate that pathogenic post-zygotic variants in breast cancer-associated genes are significantly more prevalent in normal mammary tissues of patients with adverse outcomes, such as recurrence or metastasis, compared to those without specific prognosis criteria or control individuals^15^. Monitoring these patients for nearly a decade after diagnosis allowed for a direct association between these variants and clinical outcomes. These alterations were strongly linked to disease progression, particularly recurrence, suggesting an increased risk of aggressive cancer before clinical symptoms appear. This highlights the need for expanded genetic screening and enhanced surveillance to improve personalized breast cancer management, particularly for patients with poor prognoses.

## Abbreviations

BCAP: Breast Cancer Adverse Prognoses
BCS: Breast-conserving surgery
BCUS: Breast Cancer Un-Selected
BL: Whole Peripheral Blood
PT: Primary Tumor
RM: Reduction Mammoplasty
SK: Skin
UM: Uninvolved mammary gland
UMD: Distal uninvolved mammary gland
WES: Whole Exome Sequencing

## Authors Contributions

Study design and conception: A.P., M.A.; Clinical data acquisition and interpretation: M.D., M.N., J.J., D.H.Z., J.S., E.S., M.L-J., J.H., D.B., M. Jankowski., W.Z., Ł.S., T.N., N.F., A.M., K.C., K.D-C., U. Ł., K.D., H.D., B. B-O.; Experiments: M.A., K.C., M. Koszyński, M. Jaśkiewicz., U. Ł., K.D., H.D., B. B-O., A.K.; Methodology: I.T-B.; Data analysis and interpretation: A.P., M.A., M. Koczkowska, K.C., P.G.B., P.M., M.H., M. Jąkalski, J.M.; Funding acquisition: A.P., J.P.D., K.C., I.T-B.; Visualization: M.A., M. Jąkalski; Manuscript writing – original: M.A., A.P.; Manuscript writing – review & editing: M.A., A.P., J.P.D., N.F., M. Koczkowska, M.H, P.G.B., I.T-B., A.K; Supervision: A.P., J.P.D., M. Koczkowska. All authors have read and agreed to the published version of the manuscript.

## Ethics approval and consent to participate

Tissue samples and patient histories were provided for this study by the Oncology Centre in Bydgoszcz, Jagiellonian University Hospital in Cracow, and the University Clinical Centre in Gdańsk, who, under a research protocol approved by the Bioethical Committee at the Collegium Medicum, Nicolaus Copernicus University in Toruń (approval number KB509/2010) and by the Independent Bioethics Committee for Research at the Medical University of Gdańsk (approval number NKBBN/564/2018 with multiple amendments), recruited and enrolled all donors under informed and written consent, collected, and stored all tissue samples.

## Consent for publication

Not applicable.

## Data availability

Raw duplex sequencing and WES data are available upon request in the EGA archive, under study IDs EGAS50000000538 and EGAS50000000539 respectively.

## Corresponding author

Correspondence and requests for materials should be addressed to Arkadiusz Piotrowski.

## Acknowledgments

The authors wish to thank Agata Wojdak for her help with administrative activities. We also would like to thank all the patients and volunteer control individuals for acceptance to participate in the study and sample contribution; hospital staff involved in the patient recruitment process in Oncology Center - Prof. Franciszek Łukaszczyk Memorial Hospital in Bydgoszcz, University Clinical Centre in Gdańsk and University Hospital in Cracow. We thank Dr. Leszek Kalinowski for the access to selected laboratory facilities. Figure 1 and Supplementary Figure 1 were partly generated using Servier Medical Art, licensed under a Creative Commons Attribution 3.0 Unported License (https://creativecommons.org/licenses/by/3.0/).

