## Supplementary Material for "Beyond Tumors: Reduced survival linked to pathogenic *PIK3CA* and *TP53* post-zygotic variants in the uninvolved breast tissue of recurrent cancer patients"

- Supplementary Figures
- Supplementary Tables' legends
- Supplementary References

Supplementary Figure 1

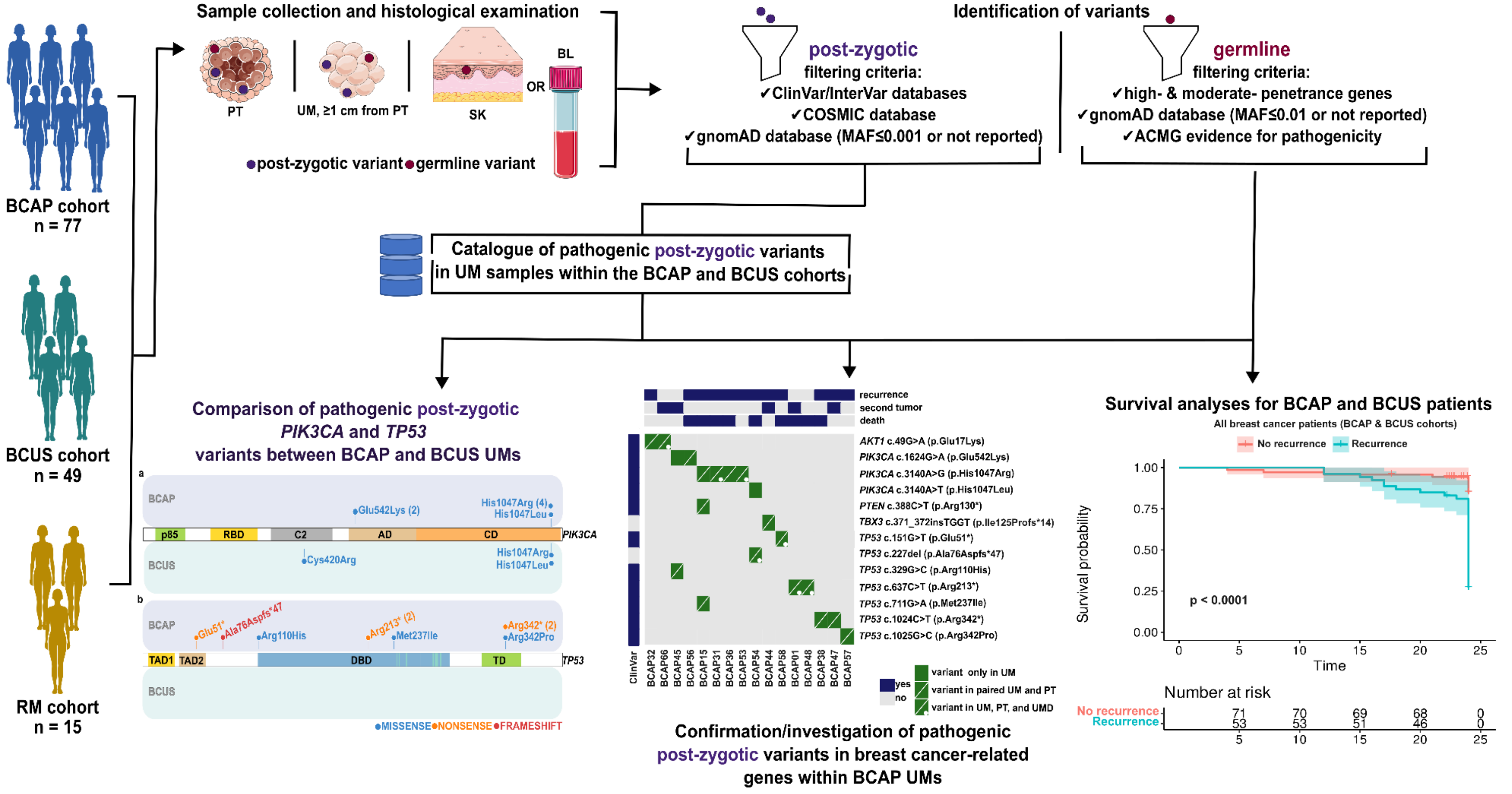


**Supplementary Figure 1. Graphical representation of the project workflow.** Three hundred seventy-eight fresh-frozen samples, including primary tumor (PT), uninvolved mammary gland (UM, ≥1 cm from PT), skin (SK), and whole peripheral blood (BL) were collected from 126 individuals diagnosed with reportedly sporadic breast cancer. Control mammary gland and BL samples were also collected from 15 individuals who underwent reduction mammoplasty surgeries, without a history of cancer (Reduction Mammoplasty, RM cohort). Breast cancer patients were stratified into two cohorts based on prognosis: those with adverse prognoses within the first 10 years post-surgery (Breast Cancer Adverse Prognoses, BCAP cohort) and those without specific prognosis-related criteria (Breast Cancer Un-Selected, BCUS cohort). For 7 BCAP individuals, additional distal UM samples (UMD, 1.5-3 cm from PT, median 2.35 cm) were also collected raising the total number of samples to 415. Pathologists confirmed normal histology of UM, UMD, and SK samples. DNA was extracted and Whole Exome Sequencing (WES) was performed to identify post-zygotic variants in matched UM and PT samples according to study criteria (Supplementary Table 2). BL or SK samples, if BL was not available, were used to distinguish between post-zygotic and germline variants. Post-zygotic variants were identified as those that were absent from the reference samples. Post-zygotic and germline variants were filtered according to study criteria (detailed in Materials and Methods). Pathogenic post-zygotic variants in breast cancer-related genes, identified within BCAP and BCUS UM samples were compiled into a single list (Supplementary Table 5), and *PIK3CA* and *TP53* variants were compared between cancer cohorts (Figure 2). Further validation of pathogenic post-zygotic variants in *AKT1*, *PIK3CA*, *PTEN*, *TBX3*, and *TP53* genes was conducted in paired UM and PT samples of the BCAP cohort via Sanger sequencing, High-Resolution Melting, or Duplex sequencing. Selected *AKT1*, *PIK3CA*, and *TP53* variants were also identified in the UMD samples of 7 BCAP patients. Hazard ratios were calculated, and Kaplan-Meier curves were generated to estimate and visualize survival probabilities over time for different patient groups.

Supplementary Figure 2

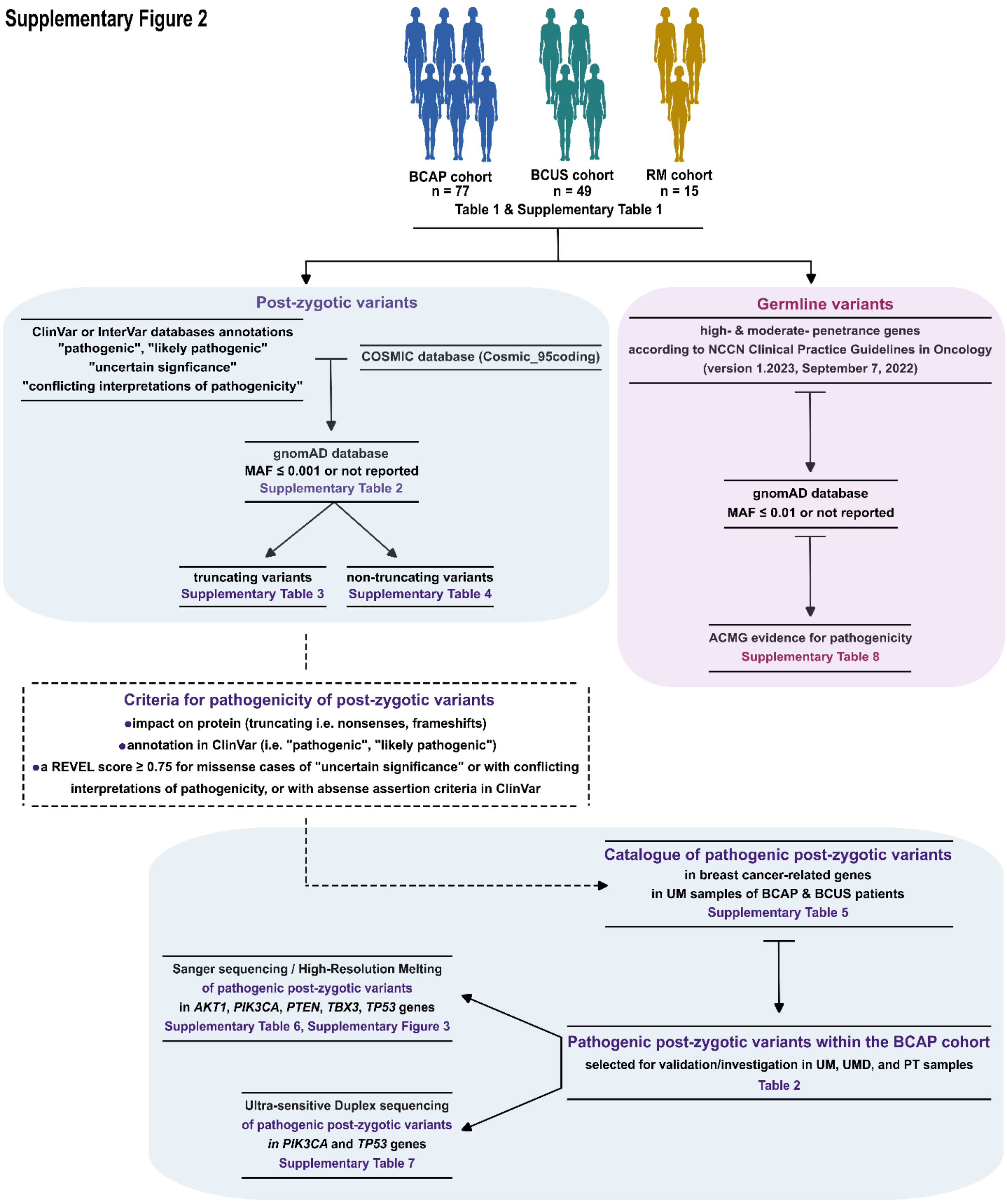


**Supplementary Figure 2. Filtering of post-zygotic and germline variants in the Breast Cancer Adverse Prognosis (BCAP), Breast Cancer Un-Selected (BCUS), and Reduction**

**Mammoplasty (RM) cohorts.** Various filtering strategies were applied to identify breast cancer-related post-zygotic and germline variants across 378 samples (UM, PT, BL, or SK) from 126 breast cancer patients and 15 controls (RM cohort) (see Materials and Methods). Detailed clinicopathological information for the BCAP and BCUS cohorts is summarized in Table 1 and Supplementary Table 1. Post-zygotic variants annotated as "pathogenic," "likely pathogenic," "uncertain significance," or "conflicting interpretations of pathogenicity" in the ClinVar or InterVar databases, as well as those listed in COSMIC (Cosmic_95coding), were included. Variants with a minor allele frequency (MAF) ≤ 0.001 or absent in gnomAD (v2.1.1) were retained for further analysis (Supplementary Table 2). Variants were categorized based on their impact on protein function: truncating variants (Supplementary Table 3) and non-truncating variants (Supplementary Table 4). Truncating variants were considered pathogenic regardless of ClinVar or InterVar annotations, while missense variants underwent *in-silico* analysis using REVEL[1] (score threshold ≥ 0.75). Variants classified as pathogenic or likely pathogenic in ClinVar, or missense variants with conflicting interpretations or without assertion criteria but supported by REVEL (≥ 0.75), were deemed pathogenic. ClinVar annotations were prioritized when selecting variants affecting breast cancer-related genes (Supplementary Table 5). Truncating variants in tumor suppressor genes were also included, even if absent in ClinVar. Selected BCAP UM, PT, and SK samples underwent further analysis using Sanger sequencing/High-Resolution Melting (Supplementary Table 6, Supplementary Figure 3) or ultra-sensitive Duplex sequencing (Supplementary Table 7). Distal UM samples (UMD) were included for seven BCAP patients. For germline variants, high- and moderate-penetrance breast cancer susceptibility genes were analyzed based on NCCN Clinical Practice Guidelines[2] (Version 1.2023, September 7, 2022). Variants with a MAF ≤ 0.01 or absent in gnomAD were subjected to *in-silico* analysis using REVEL (threshold ≥ 0.75). Germline variants were classified per the American College of Medical Genetics and Genomics and the Association for Molecular Pathology[3] guidelines. *BRCA1* and *BRCA2* variants were evaluated according to the ENIGMA BRCA1/2 Variant Curation Expert Panel (Version 1.1.0) (Clinical Genome Resource, https://www.clinicalgenome.org/affiliation/50087/, https://cspec.genome.network/cspec/ui/svi/doc/GN092, https://cspec.genome.network/cspec/ui/svi/doc/GN097). Full descriptions of germline variants in BCAP, BCUS, and RM cohorts are in Supplementary Table 8.

Supplementary Figure 3

a

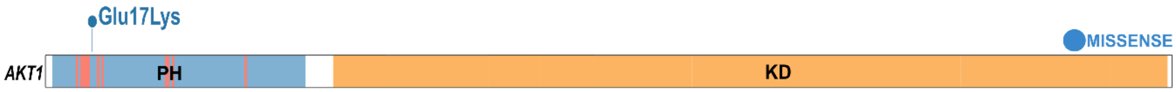


*AKT1* c.49G>A (p.Glu17Lys)

Patient ID:BCAP32

chr14:104,780,195-104,780,232

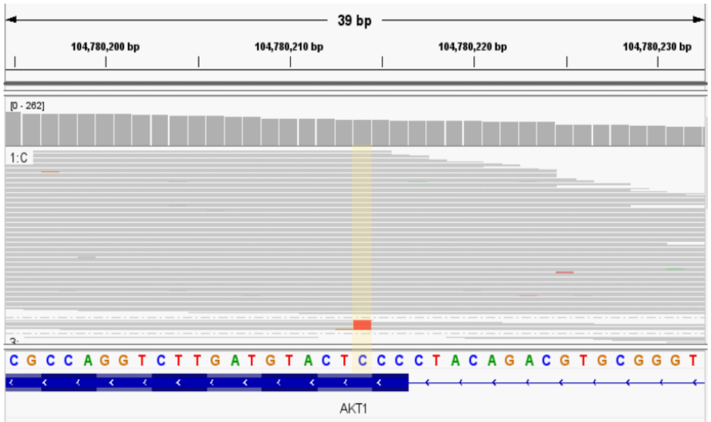


Sanger sequencing

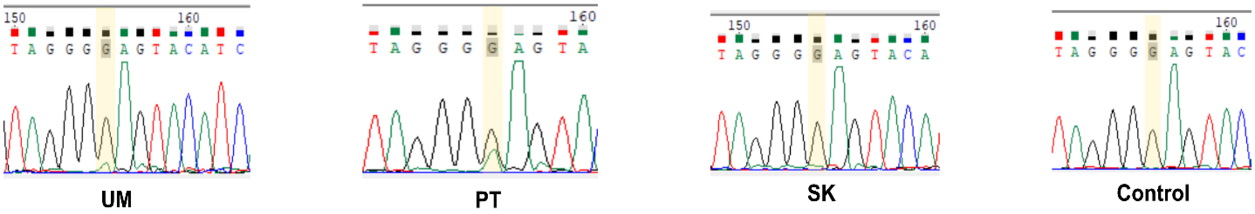


*AKT1* c.49G>A (p.Glu17Lys)

Patient ID:BCAP66

chr14:104,780,195-104,780,232

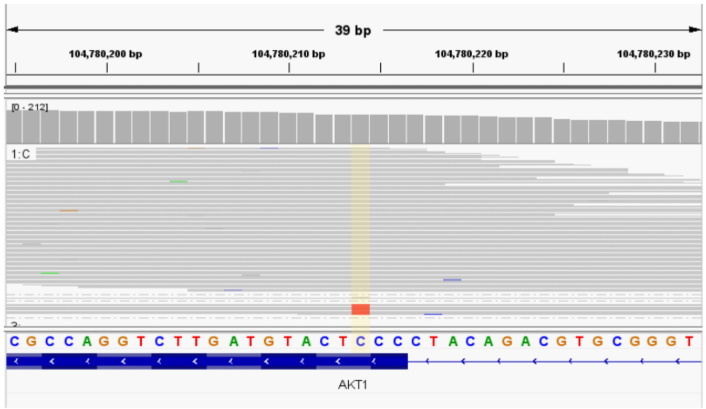


Sanger sequencing

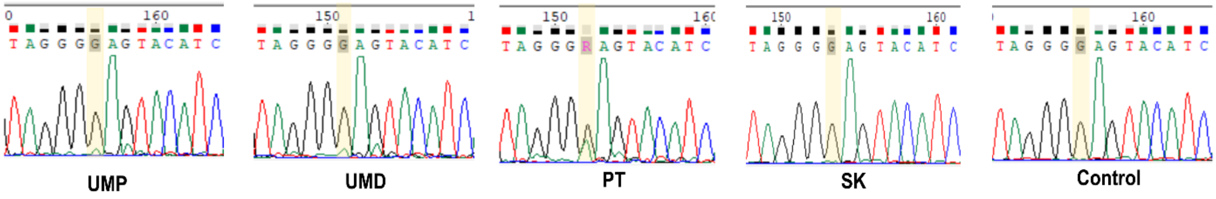

### Supplementary Figure 3

**b**

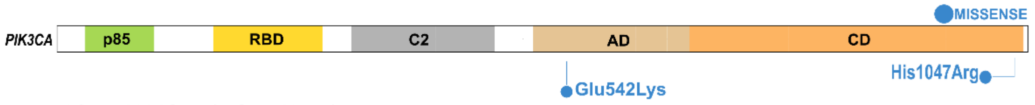


***PIK3CA* c.1624G>A (p.Glu542Lys)**

Patient ID:BCAP56

chr3:179,218,271-179,218,315

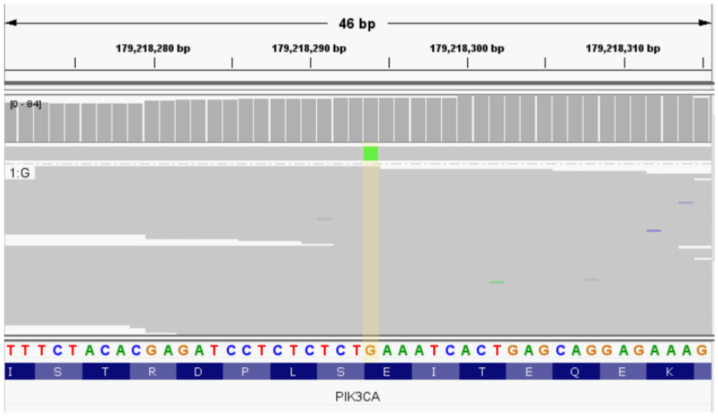


**Sanger sequencing**

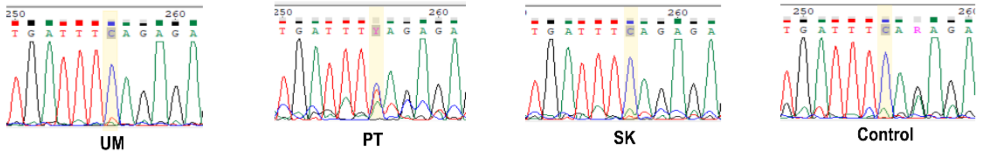


***PIK3CA* c.1624G>A (p.Glu542Lys)**

Patient ID:BCAP45

chr3:179,218,271-179,218,315

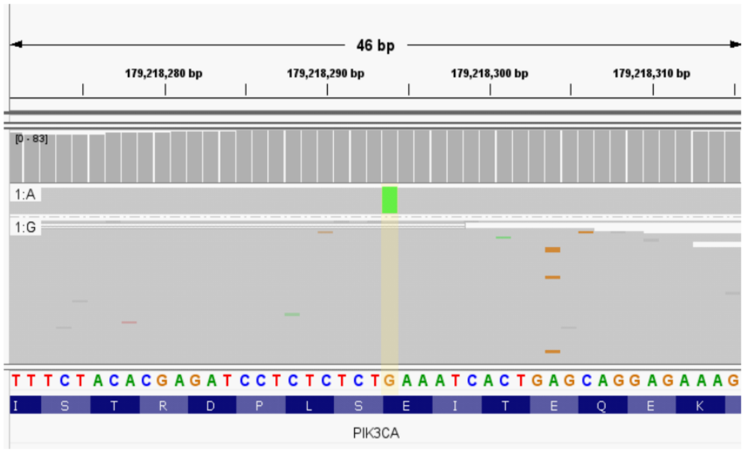


**High Resolution Melting**

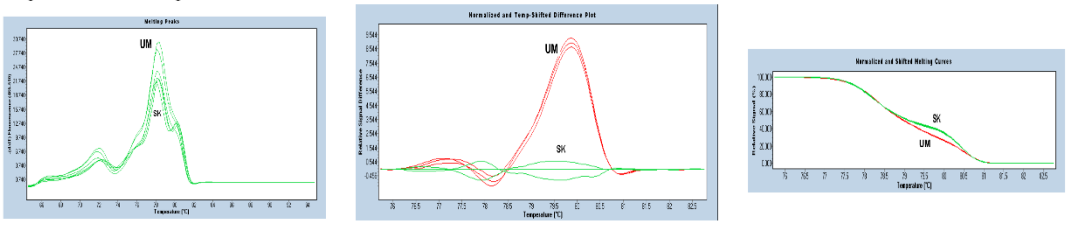

### Supplementary Figure 3

**b**

*PIK3CA* c.3140A>G (p.His1047Arg)

Patient ID:BCAP15

chr3:179,234,274-179,234,318

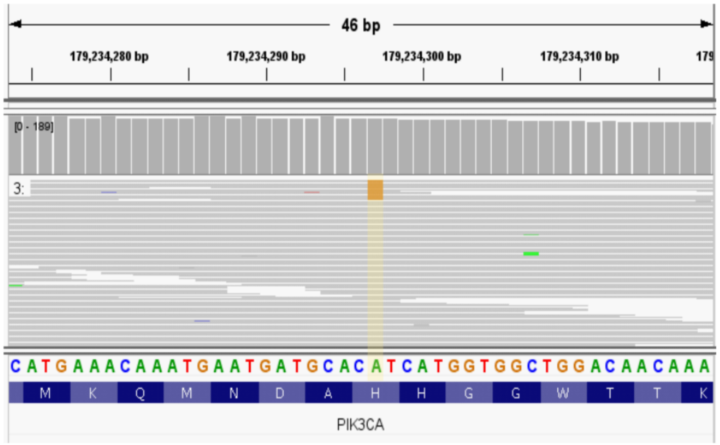


Sanger sequencing

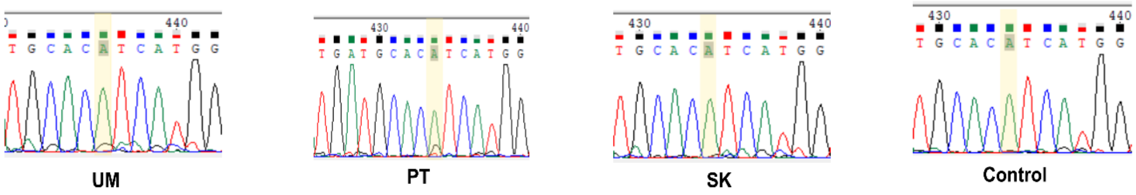


*PIK3CA* c.3140A>G (p.His1047Arg)

Patient ID:BCAP31

chr3:179,234,274-179,234,318

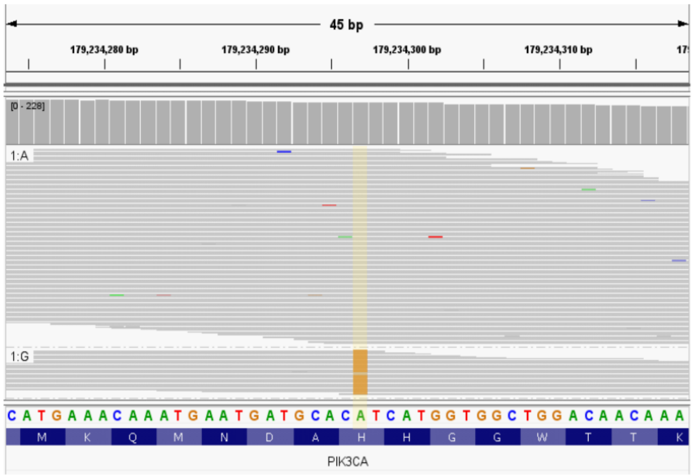


Sanger sequencing

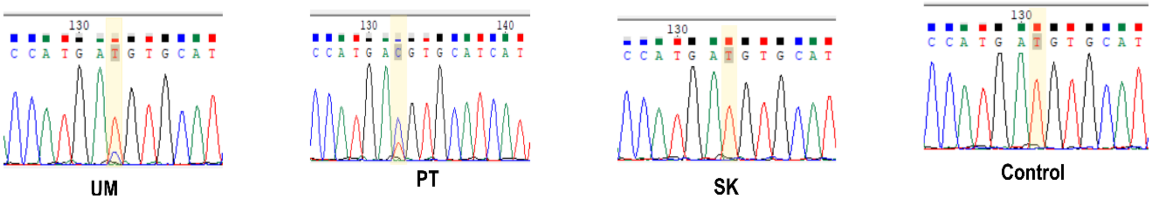

### Supplementary Figure 3

**b**

***PIK3CA* c.3140A>G (p.His1047Arg)**

**Patient ID:BCAP53**

chr3:179,234,274-179,234,318

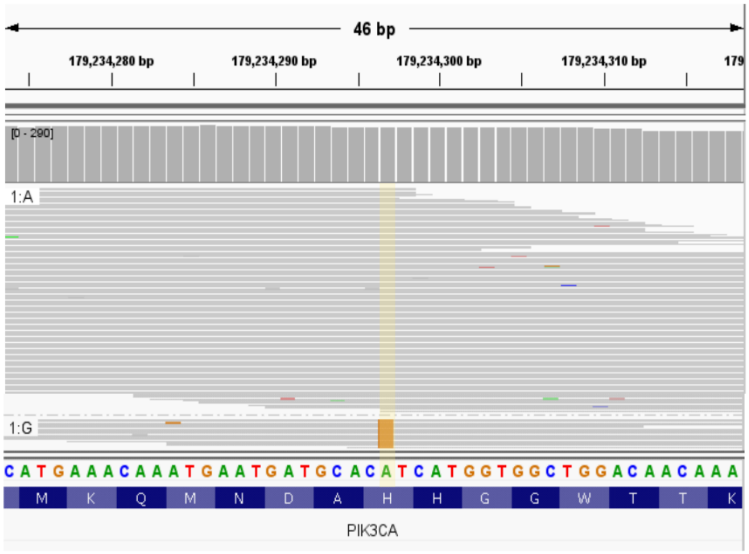


High Resolution Melting

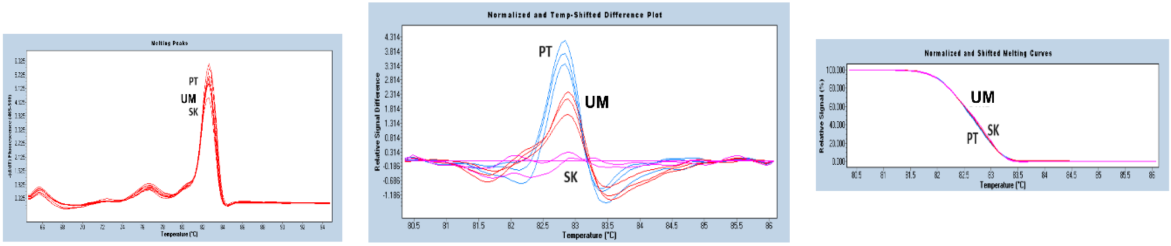

Supplementary Figure 3

c

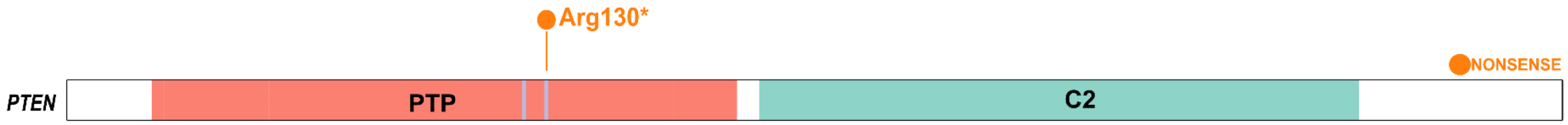


*PTEN* c.388C>T (p.Arg130*)

Patient ID:BCAP15

chr10:87,933,127-87,933,166

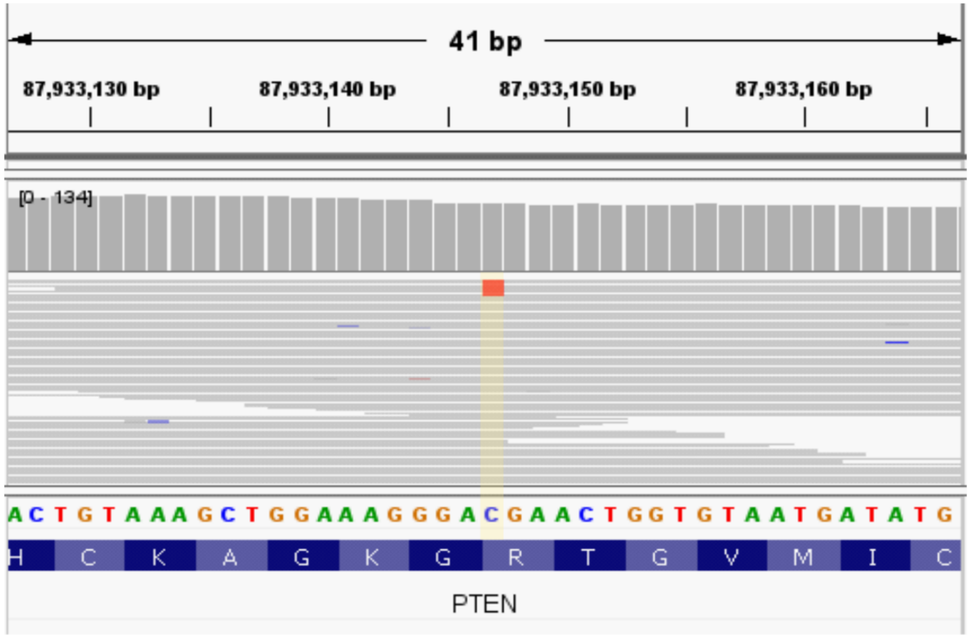


### High Resolution Melting

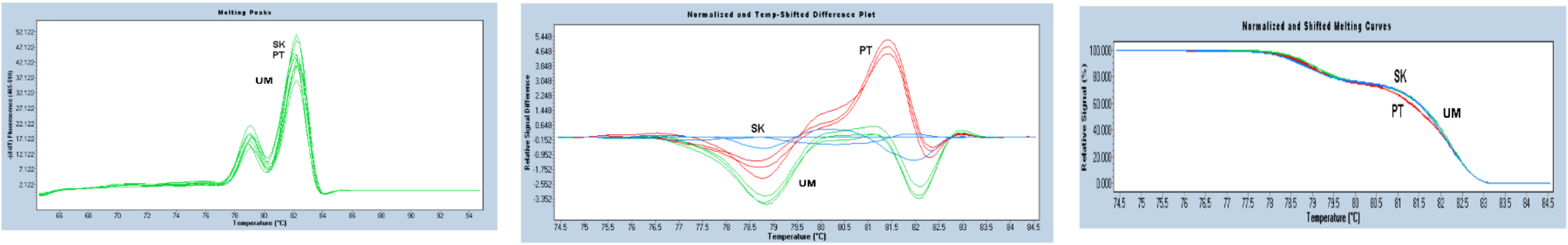

Supplementary Figure 3

d

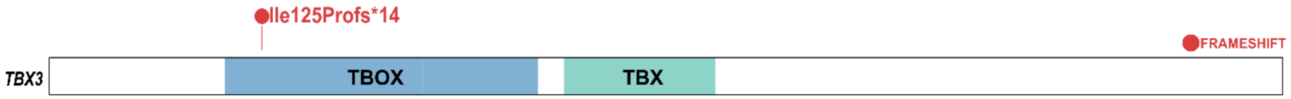


***TBX3*** **c.371_372insTGGT (p.Ile125Profs*14)**

Patient ID:BCAP44

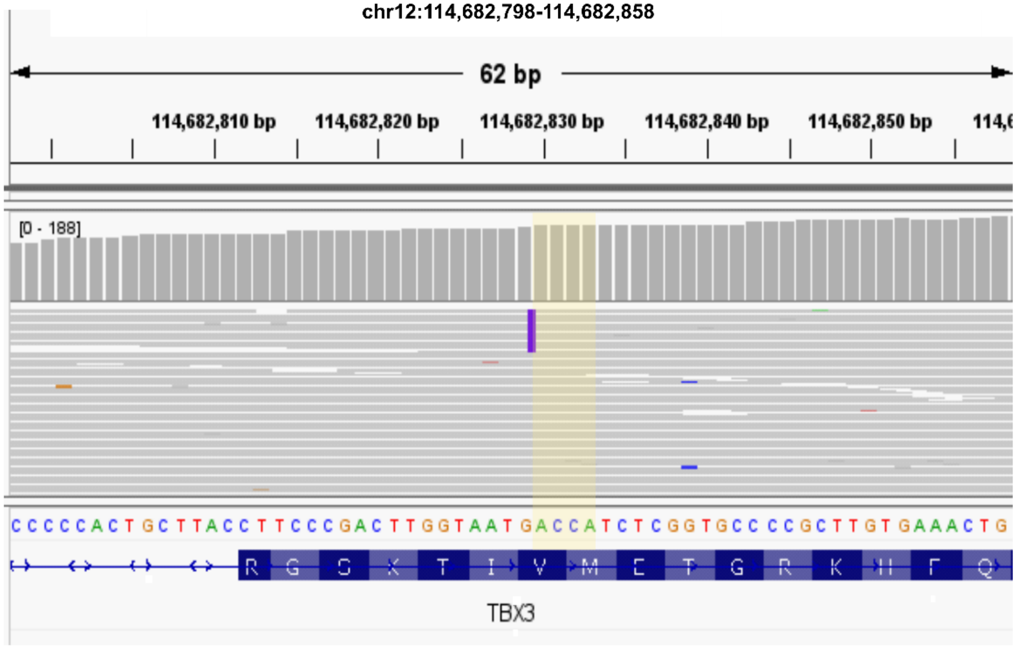


**High Resolution Melting**

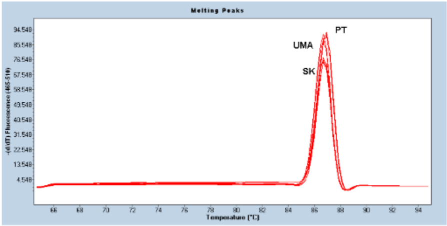


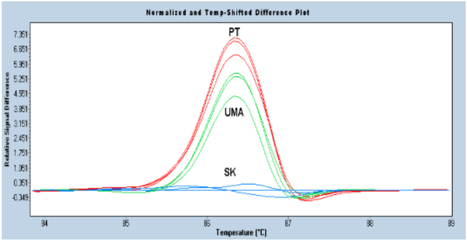


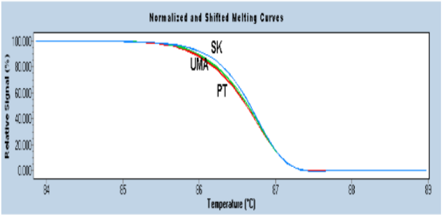

Supplementary Figure 3

e

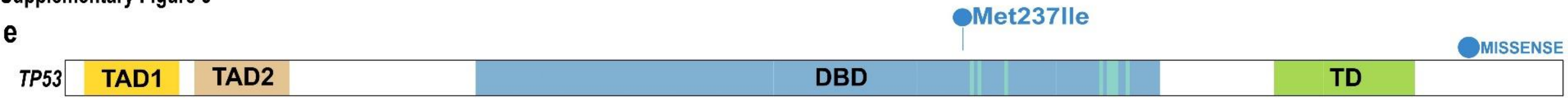


*TP53* c.711G>A (p.Met237Ile)

Patient ID:BCAP15

chr17:7,674,232-7,674,271

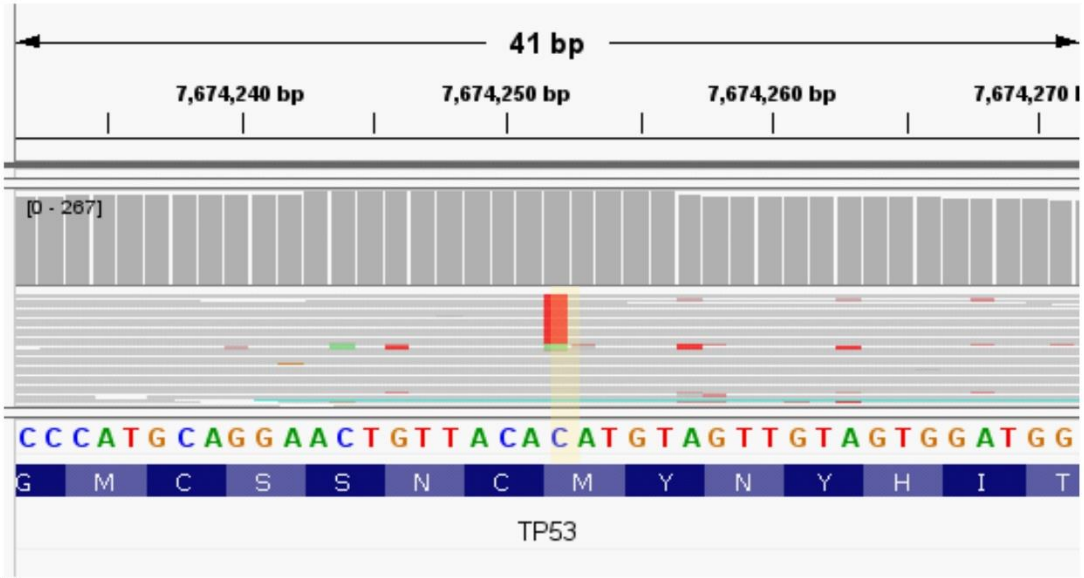


High Resolution Melting

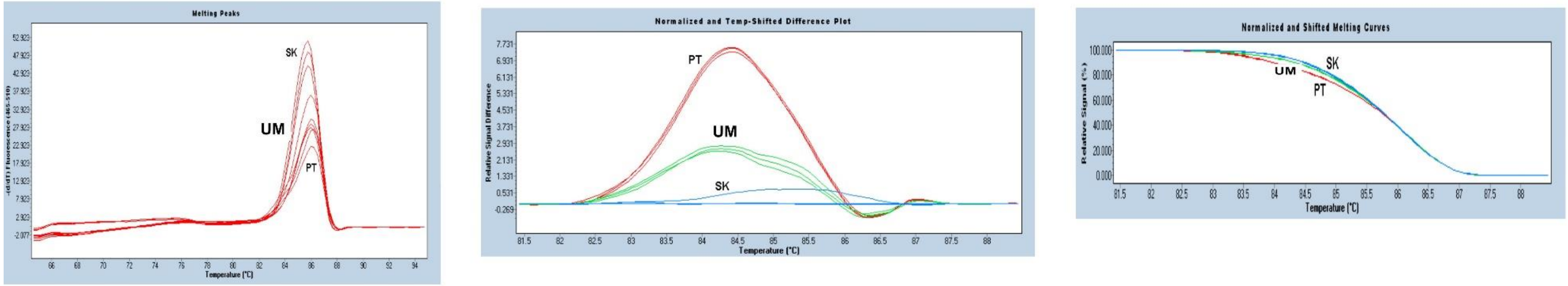

**Supplementary Figure 3. Post-zygotic DNA variants detected in the normal mammary gland samples of breast cancer patients with adverse outcomes.** The upper part of the lollipop plots presents predicted amino acid change caused by the detected variant. Missense variants are represented by blue dots, frameshift variants by red dots, and nonsense by orange. Lollipop plots were prepared based on images generated with the ProteinPaint application[4]. Middle panels present aligned reads from whole exome sequencing of UM samples with marked variant reads and were prepared based on IGV (Integrative Genomics Viewer, http://www.broadinstitute.org/igv). Lower panels include variant confirmation by Sanger sequencing or High-Resolution Melting (HRM). UM - uninvolved mammary gland, UMD - distal uninvolved mammary gland, PT - primary tumor, SK - skin, Control - peripheral blood from an unrelated individual. **a) Presentation of *AKT1* c.49G>A variant**. PH - Protein Kinase B-like pleckstrin homology domain, red lines within the PH domain indicate phosphoinositide binding sites; KD - Catalytic domain of the Serine/Threonine Kinase. **b) Presentation of *PIK3CA* c.1624G>A and c.3140A>G variants**. p85 - p85-binding domain; RBD - Ras-binding domain; C2 - C2 domain; AD - accessory domain; CD - catalytic domain. **c) Presentation of *PTEN* c.388C>T variant.** PTP - protein tyrosine phosphatase-like catalytic domain of phosphatase and tensin homolog, purple lines indicate active catalytic sites; C2 - C2 domain of PTEN tumor suppressor gene). **d) Presentation of *TBX3* c.371_372insTGGT variant**. DBD - DNA-binding domain; TBOX - T-box transcription factor domain. **e) Presentation of *TP53* c.711G>A variant.** TAD1, TAD2 - transcription activation domain 1 and 2; DBD - DNA-binding domain, DNA-binding sites are marked with red lines; TD - tetramerization domain.

**Supplementary Figure 4**

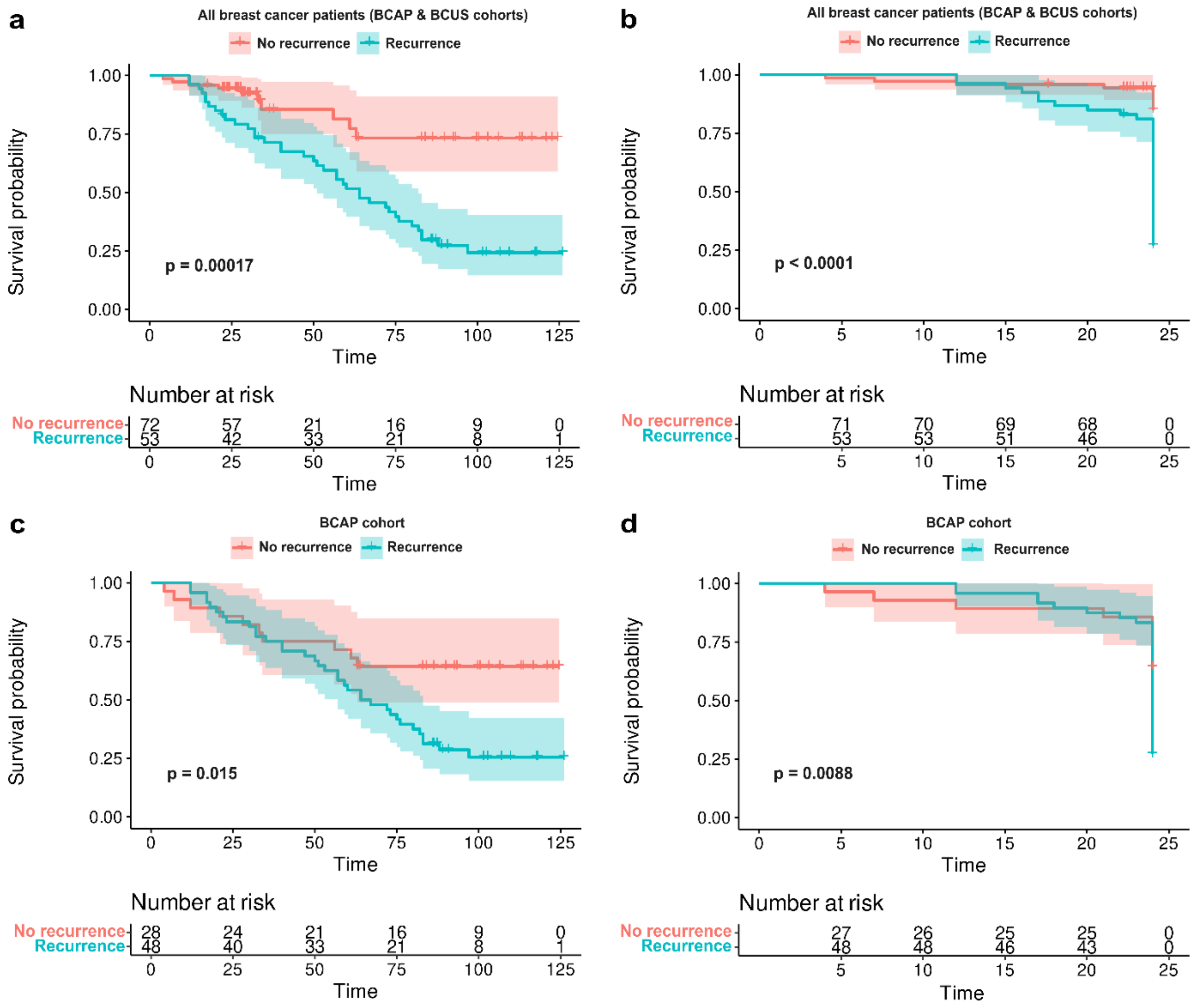


**Supplementary Figure 4. Kaplan-Meier survival curves for breast cancer patients with and without recurrent disease.** Breast cancer patients with adverse prognoses (BCAP cohort) and breast cancer patients selected without any prognoses bias (BCUS cohort) with recurrent disease had significantly lower survival probabilities compared to those without for **a) the full follow-up period** (10 and 2 years for the BCAP and BCUS cohorts respectively) (log-rank test, p=0.00017) and for **b) the first 24 months post-diagnosis** (log-rank test, p<0.0001). Within the BCAP cohort, the results were similar: **c) for the full follow-up time** (log-rank test, p=0.015) and **d) for the first 24 months after diagnosis** (log-rank test, p=0.0088). The x-axis represents time in months, and the y-axis represents the probability of survival. Censoring events are indicated by vertical ticks on the curves.

### Supplementary Tables

**Supplementary Table 1. Clinicopathological characteristics of reportedly sporadic breast cancer patients included in the Breast Cancer Adverse Prognoses (BCAP) and the Breast Cancer Un-Selected (BCUS) cohorts.** Sets of primary tumor (PT), uninvolved mammary gland (UM), and reference (whole peripheral blood, BL or skin, SK) samples were collected from 126 individuals diagnosed with reportedly sporadic breast cancer. Breast cancer patients were selected according to prognoses, i.e. patients who experienced either recurrent disease, such as local recurrence/metastasis to the breast or secondary organs (n=40), developed a second independent tumor (n=18), or both (n=8), and/or succumbed to the disease (n=45) within the following 10 years (BCAP cohort, n=77), or were recruited without prognoses-related criteria (BCUS cohort, n=49). [a]Cancer TNM stage and [b]tumor grade were assessed according to the American Joint Committee on Cancer guidelines 7th edition[5] (effective from 2010), and 8th edition[6] (effective from 2017) [c]Histological types of breast tumors recognized: invasive ductal carcinoma (IDC), invasive lobular carcinoma (ILC), neuroendocrine, mucinous, and papillary carcinoma. Comedo refers to comedocarcinoma, a type of pre-invasive breast neoplasia. [d]In situ: ductal carcinoma in situ (DCIS), lobular carcinoma in situ (LCIS). [e]Estrogen receptor (ER), progesterone receptor (PR), and Ki67 proliferation markers scores were used to assign tumors to [f]biological subtypes. [g]BCT-breast-conserving therapy; SLND - sentinel lymph node dissection; MRM (Patey) - Modified Radical Mastectomy (Patey); Q - breast quadrantectomy (included in Breast-conserving therapy procedures); ALND - axillary lymph node dissection; LS - lymphoscintigraphy; SM – Simple Mastectomy. UMD – distal UM sample (Materials and Methods). * The patient died of non-oncological causes. n.a. – not available.

**Supplementary Table 2. List of post-zygotic variants that fulfilled the study's criteria, observed within uninvolved mammary gland (UM) samples across all individuals under investigation, stratified across three distinct cohorts**. Patients diagnosed with breast cancer were stratified into either the Breast Cancer Adverse Prognoses (BCAP) or the Breast Cancer Un-Selected (BCUS) cohort, based on the presence/absence of criteria related to prognoses, respectively. Individuals, surgically treated for non-cancer reasons comprised the Reduction Mammoplasty (RM) cohort. [a]Genomic position provided for the hg38 assembly. [b]Variant annotation provided for the basic isoform of the transcript. [c]Variant annotation within the ClinVar database (https://www.ncbi.nlm.nih.gov/clinvar/). [d]Variant annotation within the InterVar database (https://wintervar.wglab.org/). [e]Variant ID in the Catalogue of Somatic Variants in Cancer database (Cosmic_95database, https://cancer.sanger.ac.uk/cosmic). [f]rsID in dbSNP build 150. [g]Variant allele frequency reported in the Genome Aggregation Database (gnomADv2.1.1; https://gnomad.broadinstitute.org/). [h]Allele frequency of detected post-zygotic variants in studied tissues: PT - primary tumor; UM - uninvolved margin. ANNOVAR Software was used for gene-based annotation (accessed between 06.2022-08.2022). n.a. – not available.

**Supplementary Table 3. List of truncating post-zygotic variants that fulfilled the study's criteria, observed within uninvolved mammary gland (UM) samples of patients included in the Breast Cancer Adverse Prognoses (BCAP) cohort.** Truncating post-zygotic variants were deemed pathogenic, irrespective of their annotation in ClinVar and InterVar databases. [a]Genomic position provided for the hg38 assembly. [b]Variant annotation provided for the basic isoform of the transcript. [c]Variant annotation within the ClinVar database (https://www.ncbi.nlm.nih.gov/clinvar/). [d]Variant annotation within the InterVar database (https://wintervar.wglab.org/). [e]Variant ID in the Catalogue of Somatic Mutations in Cancer database (Cosmic_95database, https://cancer.sanger.ac.uk/cosmic). [f]rsID in dbSNP build 150. [g]Variant allele frequency reported in the Genome Aggregation Database (gnomADv2.1.1; https://gnomad.broadinstitute.org/). [h]Allele frequency of detected post-zygotic variants in studied tissues: PT - primary tumor; UM - uninvolved margin. ANNOVAR Software was used for gene-based annotation (accessed between 06.2022-08.2022). n.a. – not available.

**Supplementary Table 4. List of non-truncating post-zygotic variants that fulfilled the study's criteria observed within uninvolved mammary gland (UM) tissue across all individuals under investigation, stratified across three distinct cohorts**. Patients diagnosed with breast cancer were stratified into either the Breast Cancer Adverse Prognoses (BCAP) or the Breast Cancer Un-Selected (BCUS) cohort, based on the presence/absence of criteria related to prognoses, respectively. Individuals, surgically treated for non-cancer reasons comprised the Reduction Mammoplasty (RM) cohort. [a]Genomic position provided for the hg38 assembly. [b]Variant annotation provided for the basic isoform of the transcript. [c]Variant annotation within the ClinVar database (https://www.ncbi.nlm.nih.gov/clinvar/). [d]Variant annotation within the InterVar database (https://wintervar.wglab.org/). [e]Variant ID in the Catalogue of Somatic Mutations in

Cancer database (Cosmic_95database, https://cancer.sanger.ac.uk/cosmic). [f]rsID in dbSNP build 150. [g]Variant allele frequency reported in the Genome Aggregation Database (gnomADv2.1.1; https://gnomad.broadinstitute.org/). [h]Variant allele frequency reported based on 1000 Genomes Project (https://www.internationalgenome.org/data). [i]Rare Exome Variant Ensemble Learner (REVEL) score[1]. [j]Pathogenicity classification according to Mendelian Clinically Applicable Pathogenicity (M-CAP) Score: (D) - damaging, (T) - tolerated[7]. [k]Sorting Intolerant from Tolerant (SIFT) score: (D) - damaging, (T) – tolerated[8]. [l]Polyphen2 HumDiv and Polyphen2 HumVar[9]. [m]Combined Annotation Dependent Depletion (CADD) phred-like score[10]. [n]Protein Variation Effect Analyzer (PROVEAN) phred-like score: (D) - deleterious, (N) – neutral[11]. [o]Genomic Evolutionary Rate Profiling rejected substitutions (GERP++ RS) score[12]. [p]SF, MaxEntScan, and NNSplice splice prediction algorithms were used for *in-silico* splicing analysis as implemented in Alamut Visual software v1.8.1 (September - October 2023). [q]Allele frequency of detected post-zygotic variants in studied tissues: PT - primary tumor; UM - uninvolved margin. ANNOVAR Software was used for gene-based annotation (accessed between 06.2022-08.2022). SIFT, Polyphen2HDIV, and Polyphen2HVAR were accessed via Alamut Visual software v1.8.1. (September-October 2023). 1000Genomes and CADD scores were acquired via wANNOVAR (March - June 2023). n.a. – not available.

**Supplementary Table 5. List of pathogenic post-zygotic variants, according to the study criteria, observed within uninvolved mammary gland (UM) samples across all recruited breast cancer individuals, stratified in two distinct cohorts.** Variants were declared as pathogenic based on their annotation in the ClinVar database, REVEL score ≥ 0.75 (for missense cases), and effect on encoded protein (Materials and Methods). Patients diagnosed with breast cancer were stratified into either the Breast Cancer Adverse Prognoses (BCAP) or the Breast Cancer Un-Selected (BCUS) cohort, based on the presence/absence of criteria related to prognoses, respectively. [a]Genomic position provided for the hg38 assembly. [b]Variant annotation provided for the basic isoform of the transcript. [c]Variant annotation within the ClinVar database (https://www.ncbi.nlm.nih.gov/clinvar/). [d]Variant ID in the Catalogue of Somatic Mutations in Cancer database (Cosmic_95database, https://cancer.sanger.ac.uk/cosmic). [e]rsID in dbSNP build 150. [f]Variant allele frequency reported in the Genome Aggregation Database (gnomADv2.1.1; https://gnomad.broadinstitute.org/). [g]Variant allele frequency reported based on 1000 Genomes Project (https://www.internationalgenome.org/data). [h]Rare Exome Variant Ensemble Learner

(REVEL) score[1]. [i]Pathogenicity classification according to Mendelian Clinically Applicable Pathogenicity (M-CAP) Score: (D) - damaging, (T) - tolerated[7]. [j]Sorting Intolerant from Tolerant (SIFT) score: (D) - damaging, (T) – tolerated[8]. [k]Polyphen2 HumDiv and Polyphen2 HumVar[9]. [l]Combined Annotation Dependent Depletion (CADD) phred-like score[10]. [m]Protein Variation Effect Analyzer (PROVEAN) phred-like score: (D) - deleterious, (N) – neutral[11]. [n]Genomic Evolutionary Rate Profiling rejected substitutions (GERP++ RS) score[12]. [o]SF, MaxEntScan, and NNSplice splice prediction algorithms were used for *in-silico* splicing analysis as implemented in Alamut Visual software v1.8.1 (September - October 2023). [p]Allele frequency of detected post-zygotic variants in studied tissues: PT - primary tumor; UM - uninvolved margin. ANNOVAR Software was used for gene-based annotation (accessed between 06.2022-08.2022). SIFT, Polyphen2HDIV, and Polyphen2HVAR were accessed via Alamut Visual software v1.8.1. (September - October 2023). 1000Genomes and CADD scores were acquired via wANNOVAR (March - June 2023). n.a. – not available.

**Supplementary Table 6. Pathogenic post-zygotic variants in breast cancer-associated genes observed within the uninvolved mammary gland (UM) samples of breast cancer patients included in the Breast Cancer Adverse Prognoses (BCAP) cohort and validated by Sanger sequencing or High-Resolution Melting**. Variants were initially detected in UM and primary tumor (PT) tissues with standard Whole Exome Sequencing (WES). [a]Variant annotation provided for the basic isoform of the transcript. [b]Pathogenicity classification according to the ClinVar database. [c]ID of the variant in the Cosmic_95 coding database. [d]rsIDs in dbSNP build 150. [e]Tissue allele frequency of the detected variants in matched UM and PT tissues detected with WES. Detailed description of selected post-zygotic variants is provided in Supplementary Table 5. Confirmation of post-zygotic variants by Sanger sequencing or High-Resolution Melting is available in Supplementary Figure 3. *The *AKT1* variant was also confirmed in the distal UM sample (UMD) of patient BCAP66. n.a. - not available.

**Supplementary Table 7. Pathogenic post-zygotic variants in *PIK3CA* and *TP53* genes observed within the uninvolved mammary gland (UM) samples of breast cancer patients included in the Breast Cancer Adverse Prognoses (BCAP) cohort and validated by Duplex sequencing.** Variants were initially detected in UM and primary tumor (PT) tissues with standard Whole Exome Sequencing (WES). Distal UM samples (UMD), available for 6 individuals, were included in the Duplex sequencing experiment. [a]Variant annotation provided for the basic isoform of the transcript. [b]Variant allele frequency in PT tissues, identified via WES. PT tissues were not included in the Duplex sequencing experiment. *Variants were confirmed via Sanger sequencing/High-Resolution Melting. Detailed description of selected post-zygotic variants is provided in Supplementary Table 5. n.a. – not applicable.

**Supplementary Table 8. Pathogenic germline variants in high and moderate penetrance genes for breast cancer according to NCCN Clinical Practice Guidelines in Oncology[33] (Version1.2023, September 7, 2022), detected in breast cancer patients included in the Breast Cancer Adverse Prognoses (BCAP) and the Breast Cancer Un-Selected (BCUS) cohorts.** [a]Genomic position provided for the hg38 assembly. [b]Variant annotation provided for the basic isoform of the transcript. [c]Pathogenicity classification according to the current American College of Medical Genetics and Genomics (ACMG) guidelines, classification based on the following criteria, cited directly from Richards et al.[3]. PVS1 null variant (nonsense, frameshift, canonical ±1 or 2 splice sites, initiation codon, single or multiexon deletion) in a gene where LOF is a known mechanism of disease; PM1 Located in a mutational hot spot and/or critical and well-established functional domain (e.g., active site of an enzyme) without benign variation; PM2 Absent from controls (or at extremely low frequency if recessive) in Exome Sequencing Project, 1000 Genomes Project, or Exome Aggregation Consortium; PP4 Patient's phenotype or family history is highly specific for a disease with a single genetic etiology; PP5 Reputable source recently reports variant as pathogenic, but the evidence is not available to the laboratory to perform an independent evaluation. For *BRCA1* and *BRCA2* variants, evaluation was performed according to the Evidence-based Network for the Interpretation of Germline Mutation Alleles (ENIGMA) *BRCA1* and *BRCA2* Variant Curation Expert Panel[13]. [d]Variant annotation within the ClinVar database (https://www.ncbi.nlm.nih.gov/clinvar/). [e]Variant ID in the Catalogue of Somatic Mutations in Cancer database (Cosmic_95database, https://cancer.sanger.ac.uk/cosmic). [f]rsID in dbSNP build 150. [g]Variant allele frequency reported in the Genome Aggregation Database (gnomADv2.1.1; https://gnomad.broadinstitute.org/). [h]Variant allele frequency reported based on 1000 Genomes Project (https://www.internationalgenome.org/data). [i]Number of entries and variant classification on pathogenicity according to the Leiden Open (source) Variation Database (LOVD) (https://www.lovd.nl/3.0/home) (last accessed on 18.05.2024). [j]Allele frequency of detected germline variants in studied tissues: SK – skin; BL – whole peripheral blood; PT - primary tumor;

UM - uninvolved margin. ANNOVAR Software was used for gene-based annotation (accessed between 06.2022-08.2022). 1000Genomes information was acquired via wANNOVAR (as of 21.05.2024). n.a. – not available.
